# Partnership-focussed Principles-driven Online co-Design (P-POD): a mixed-methods evaluation of a novel online co-design process

**DOI:** 10.1101/2023.05.25.23290507

**Authors:** Ms Free Coulston, Prof Alicia Spittle, Dr Cassie McDonald, Dr Rachel Toovey, Dr Kate L Cameron, Kimberley Attard, Loni Binstock, Isaac Fletcher, Adie Delaney, Tayla Murphy, Caroline Keating, Dr Kath Sellick

## Abstract

**Purpose:** Co-design involves stakeholders in designing rehabilitation interventions that impact their health (end-users) or professional lives (clinicians and researchers). Partnership-focussed Principles-driven Online co-Design (P-POD) is proposed and evaluated as an authentic approach to adapting research co-design into an online environment.

**Materials and methods:** A community-based participatory research approach scaffolded the co-design process and convergent mixed-methods evaluation. P-POD involved 10 stakeholders (parents, clinicians, coaches, and researchers) in eight 90-minute workshops to co-design a circus-based rehabilitation intervention to improve participation for preschool-aged children born preterm (premmies). P-POD was evaluated via anonymous surveys during workshops and semi-structured interviews upon completion of the process. Data were analysed using descriptive statistics and reflexive thematic analysis.

**Results:** The resulting co-designed intervention is “CirqAll: preschool circus for premmies”. Evaluation of P-POD indicated adherence to guiding principles of stakeholder involvement and co-design. Themes describe participants’ experiences of the supportive online culture, room for healthy debate, power-sharing, and multiple definitions of success.

**Conclusions:** P-POD appears to provide an authentic transition of research co-design into an online environment. P-POD was successfully used with stakeholders to produce a paediatric rehabilitation intervention, and benefits from the online approach align with, and extend on, those reported in the literature on in-person co-design approaches.

## Introduction

Stakeholder involvement in health research and intervention design is advocated for on pragmatic grounds, as it ensures that people most affected by the research can provide their unique perspective on constructs such as barriers, facilitators and outcomes, thus improving the service and its uptake [1–3]. Involving stakeholders in service and intervention design has been shown to increase the likelihood of implementation success by enhancing the relevance of services, increasing end-user satisfaction, and improving stakeholder ownership and uptake of healthcare initiatives [1,4–10]. Involvement of stakeholder expertise can also be argued for as a moral imperative whereby research that affects patients and their families should also facilitate their input as evidenced in the disability rights call to action ’nothing about us without us’ [7,8,10–14].

Stakeholders in health research may be defined as *“an individual or group who is responsible for or affected by health- and healthcare-related decisions”* [15,p. 459]. Types of stakeholders may range from the population that is the intended target of the health intervention (end-users such as families of children with disability), to service providers (such as clinicians and program delivery personnel), and further, to academic stakeholders such as researchers [15–18]. End-users (also referred to as consumers) may provide insight into their specific needs, while clinicians and other program providers (collectively termed service-based stakeholders) have unique contextual knowledge on intervention development, implementation, and referral pathways. Academic stakeholders provide evidence from current research, and expertise in research methods and methodologies [10,16–18]. Stakeholders may also bring lived and/ or professional expertise across multiple areas.

Stakeholder involvement (also known as patient and public involvement (PPI) [3]) must be considered carefully. Tokenistic or ill-defined approaches can create distrust of the research process, and further entrenches traditional problematic power distributions such as research *on* end-users, rather than *with* end-users [13,19]. The International Association for Public Participation’s IAP2 Spectrum of Public Participation provides guidance regarding different levels of stakeholder involvement, from the lowest level where stakeholders are provided with information in a top-down manner (*inform*), to *empower* which describes the highest level as stakeholder-driven research [20]. Defining research processes in accordance with this framework allows clarity for all parties regarding the level of stakeholder involvement, thus ensuring transparency and clear expectations of the process for stakeholders [20].

An example of a process that facilitates the goals of meaningful stakeholder engagement in health research, is research co-design. Co-design may be defined simply as *“**Co:** together, mutually, in common; **Design:** to prepare the plans, form and structure for a work”* [21,p.9]. Co-design is a process that brings together stakeholders to develop shared solutions in a series of workshops [22]. On the IAP2 spectrum, co-design usually aims to either *“empower”* stakeholders or *“collaborate”* with stakeholders by partnering to create solutions that meet identified needs [20]). The essential element of co-design that sets it apart from other research methods is that it must go beyond consultation, and instead, support and facilitate stakeholders to drive the design and decision-making as true collaborators, ensuring that their needs and experiences are at the centre of the process [2,6,8,20]. The problematic issues of tokenistic or ill-defined methods described above concerning stakeholder involvement have also been reported in the literature on research co-design. Often this occurs when consultation is masked as co-design, resulting in decision-making power remaining with the researchers. This may further perpetuate problematic power differentials and distrust of research co-design processes [23].

Traditionally, research co-design has been undertaken in a series of in-person workshop-style sessions, however, in recent years co-design has been challenged to move into an online environment, particularly during the COVID-19 pandemic. There is limited methodological literature describing how to authentically transition research co-design into an online environment, both to ensure that stakeholders have positive experiences engaging in an online environment and also that relevant, high-quality outcomes are produced. Exploring online methods is vital to capitalise on the convenience and accessibility that online approaches provide, and to ensure stakeholder involvement continues to enrich health intervention design regardless of COVID-19 or other restrictions [24,25]. This study describes the development and evaluation of a novel online research co-design method developed by the authors: Partnership-focussed Principles-driven Online co-Design approach (P-POD). P-POD was used in developing a circus-based rehabilitation intervention to increase physical activity participation for preschool-aged children born preterm.

Preschool-aged children (three to five years old) born very preterm participate in less physical activity and engage in more stationary behaviour than term-born children [26,27]. The benefits of participating in physical activity during childhood and its benefit to long-term health have been well established [28]. For children born prematurely, who experience increased risk in multiple health domains [29–33], engaging in adequate physical activity may be even more essential. Furthermore, physical activity levels as a child are shown to be predictive of participation as an adult [34], requiring the prioritization of participation-focused rehabilitation interventions as a vital component of health promotion for this cohort [35].

Circus is one possible intervention that could improve physical activity and participation. A scoping review by the authors [36] describes how circus activities show promise for improvements in physical and social-emotional outcomes in paediatric populations with biopsychosocial challenges including developmental delay and autism, both of which are prevalent in children born preterm. A community-based circus activity that targets physical activity participation for children born preterm is a novel intervention and applying a research co-design process to ensure that the intervention meets the needs of key stakeholders is important for successful implementation.

Evaluation of the novel online co-design process itself is essential to understand the unique challenges and benefits of P-POD [10,37,38]. Furthermore, understanding the extent to which P-POD was able to adhere to guiding principles will indicate if this online research co-design approach retains the authenticity of traditional face-to-face approaches [21]. This will inform how authentic online research co-design can be achieved and provide recommendations to capitalise on the accessibility and convenience of online approaches. Therefore, the primary objectives of this study are to:

1. Describe the novel P-POD process,
2. Evaluate the P-POD process to assess its adherence to guiding principles and to understand the participating stakeholders’ experience,
3. Describe the resulting co-designed intervention when P-POD is applied to co-design a circus-based rehabilitation intervention for children born preterm.

These objectives aim to answer the research questions: (1) Can research co-design be conducted authentically in an online environment? and; (2) Can P-POD produce a co-designed paediatric rehabilitation intervention?

## Materials and Methods

### Research design

This research sits within a community-based participatory research (CBPR) approach [16]. CBPR aims to foster partnerships between researchers and other stakeholders (particularly end-users) to co-create and translate knowledge into practice and reduce health inequities [16]. A co-design approach was used for the development of the rehabilitation intervention, and a mixed-methods design for the evaluation component.

This study received ethical approval from The Royal Children’s Hospital Human Research and Ethics Committee (Ethics approval number: HREC/15/RCHM/110).

#### Part 1: Co-designing the rehabilitation intervention

Literature on partnership-focused frameworks for stakeholder involvement and research on co-design describes the importance of guiding principles to inform engagement processes, decision-making, and design activities. To ensure an authentic evidence-based transition into an online environment, a set of guiding principles for P-POD were chosen by thematically categorising principles derived from the literature [2,13,15,17,21,22,39–42]. The final four thematic categories became P-POD’s guiding principles, and these were: being inclusive, respectful, participatory, and outcomes-focused (see Figure 1). Table 1 illustrates how each principle guided the selection of strategies and activities for the transition of the research co-design process to the online environment. P-POD was then delivered over a series of workshops held on a web conferencing platform (and other methods described in Table 1) to design the paediatric circus-based intervention.

**Figure 1:**
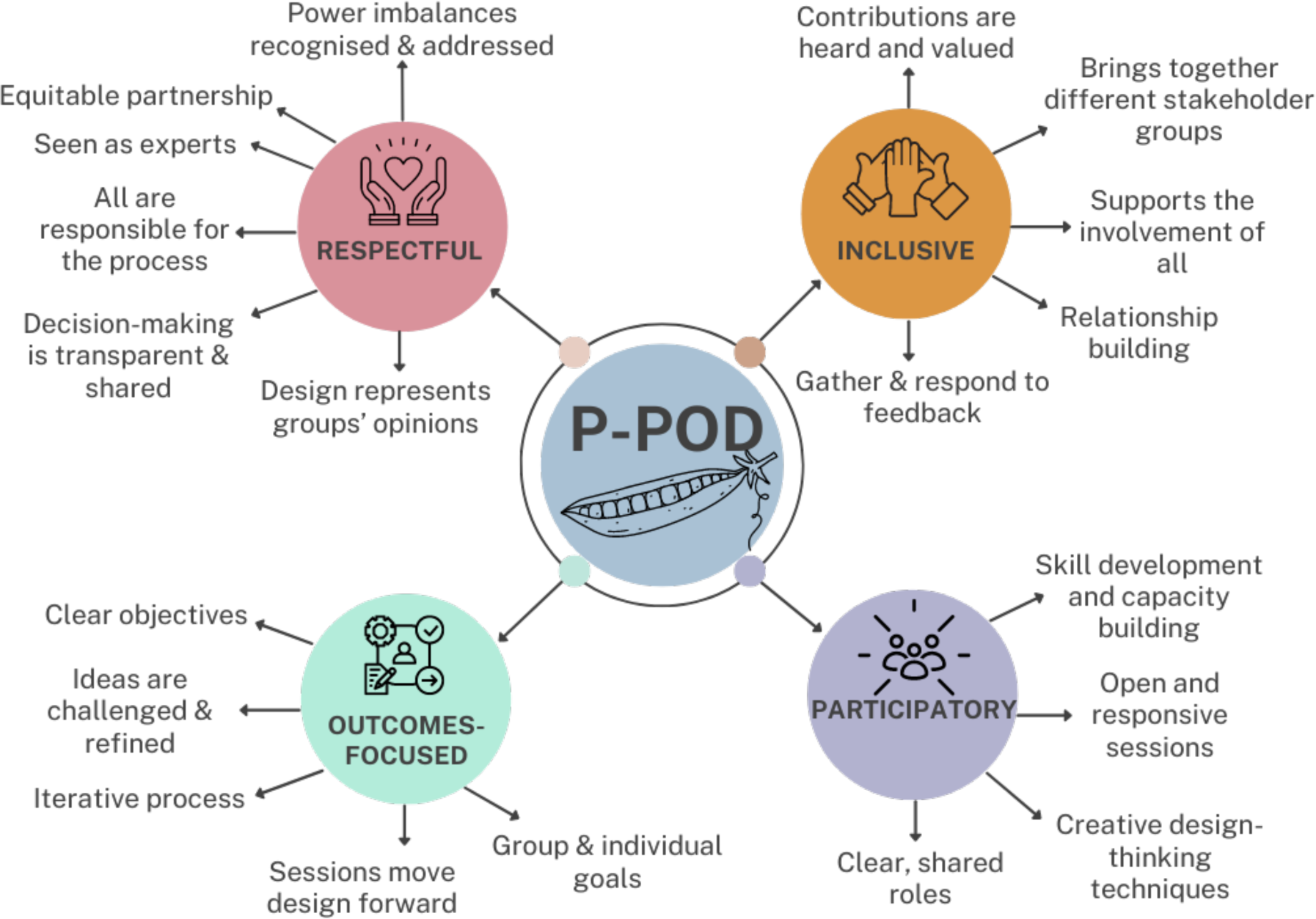
Partnership-focussed Principles-driven Online co-Design’s [P-POD] guiding principles. Figure 1 Alt Text: A mind map showing a centre circle titled P-POD, with four branches titled respectful, inclusive, participatory and outcomes-focussed respectively which list strategies employed.

**Table 1:**
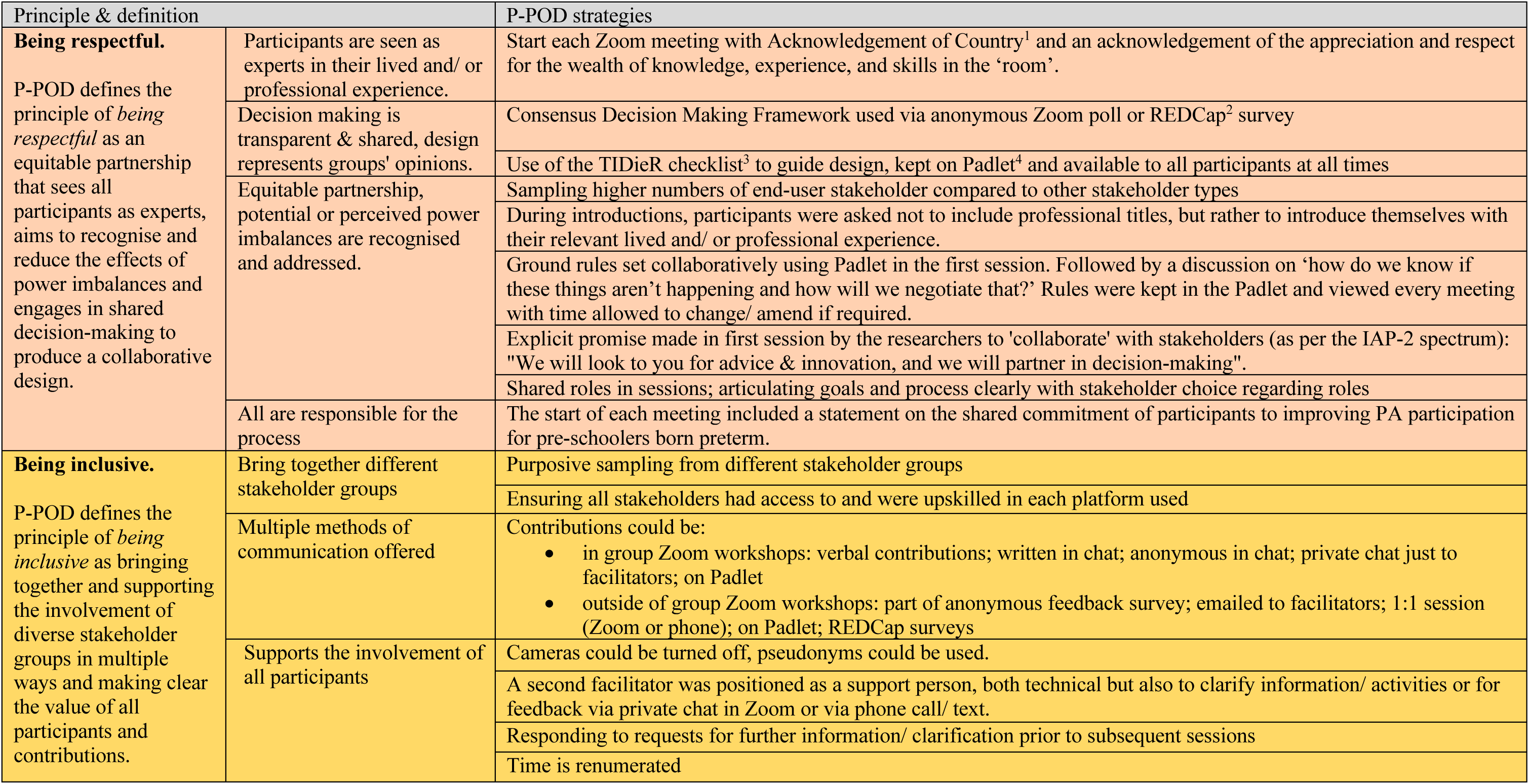
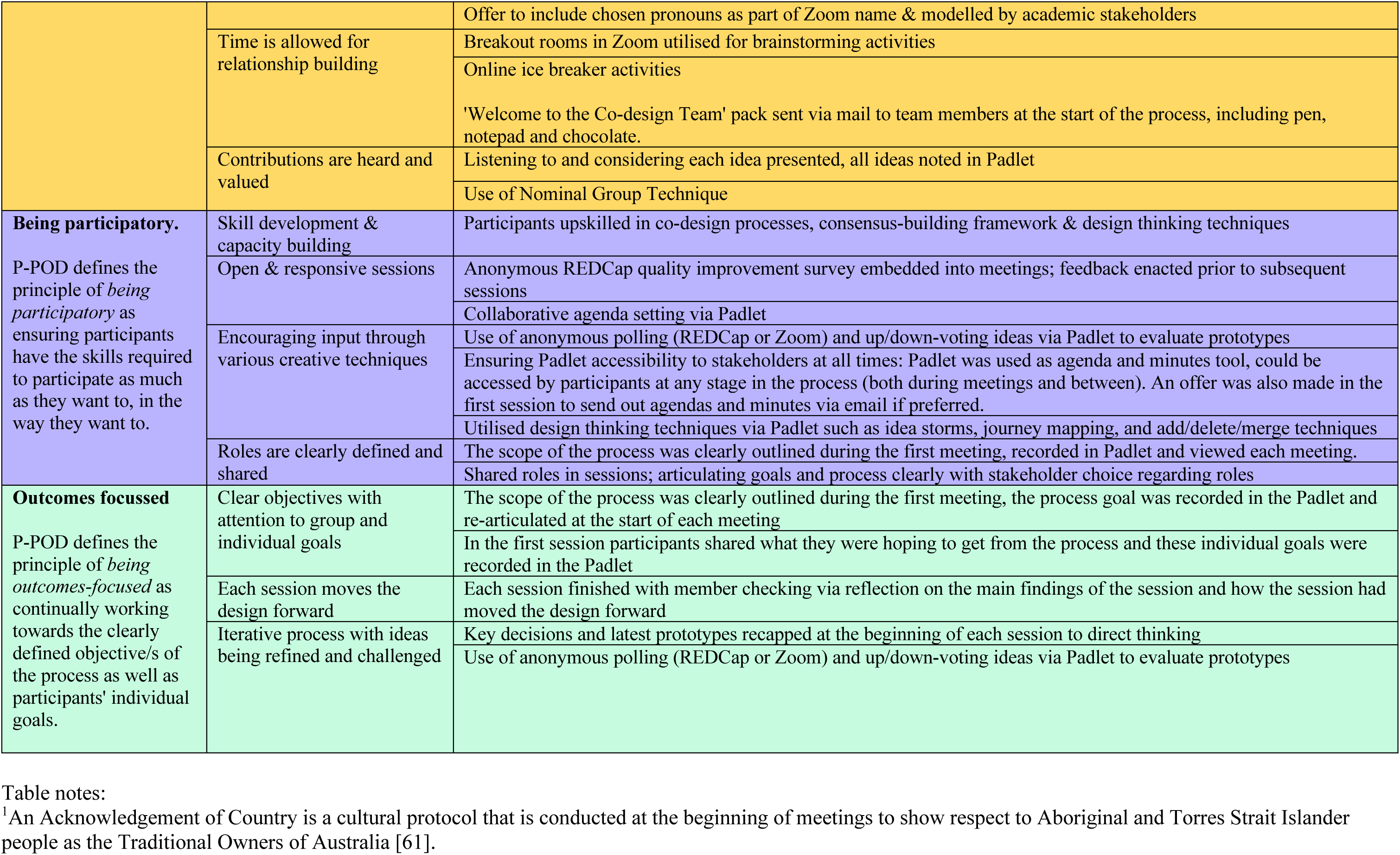
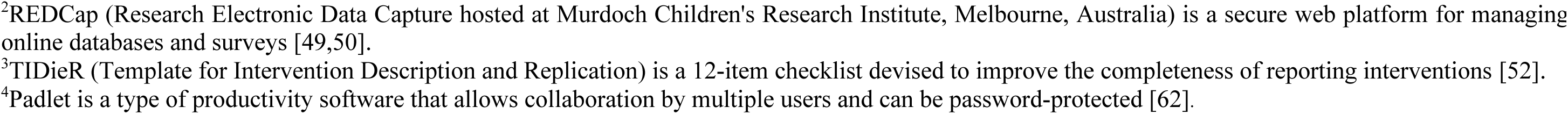
Strategies and activities utilised in Partnership-focussed Principles-driven Online co-Design [P-POD] mapped to the four guiding principles.

#### Part 2: The evaluation of P-POD

A convergent mixed-methods design using surveys and semi-structured interviews was selected for evaluation of P-POD (Figure 2). This evaluation method allowed experts in each method to provide support to the lead author in each arm of the study [43]. Bringing the data analysis together at the interpretation stage in a convergent design facilitates a depth of understanding that aligns with the recommendations in the literature for the use of mixed-methods in evaluation of co-design and stakeholder involvement processes [2,13,43].

**Figure 2:**
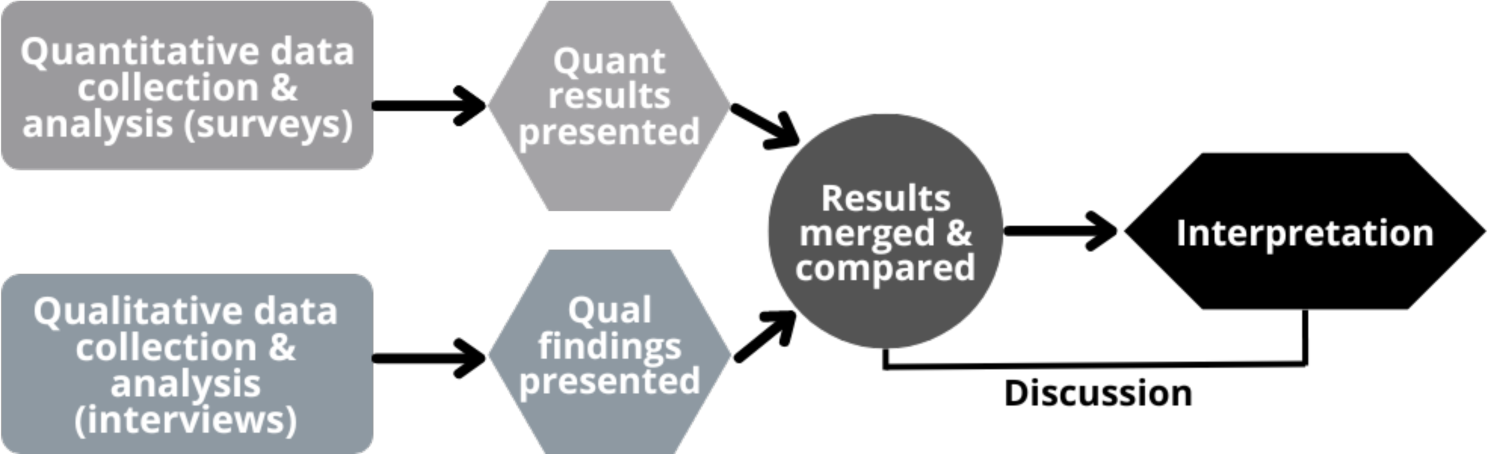
Convergent mixed-methods design employed in the evaluation of the co-design process. Figure 2 Alt Text: A flow chart showing that qualitative and quantitative data were collected and analysed separately and merged for interpretation in the discussion.

### Participants

In sampling, the aim was to consider who might be most affected by the intervention, as well as having diversity in stakeholder groups to explore all the important perspectives and types of expertise (end-users who know their context and needs, service-based stakeholders with experience of intervention delivery and referral, and academic stakeholders with relevant research evidence and practice) [13,17]. Prior to P-POD, a detailed mixed-methods study was undertaken by the authors which focussed on understanding the needs and preferences of key stakeholders with regard to their experiences with recreational physical activities for children born preterm [44]. P-POD participants (also referred to in this manuscript as “team members” or “the co-design team”) were purposively selected from participants involved in this prior study using maximum-variation sampling based on survey and interview responses. This approach aimed to capture team members with diverse experiences, opinions, (and practice settings for service-based stakeholders).

The co-design team involved in P-POD consisted of ten members: end-users (parents of preschool-aged children born extremely preterm [<28 weeks gestational age]; *n=4*), service-based stakeholders (paediatric clinicians; *n=2,* and circus coaches with expertise in the preschool-age; *n=2*), and academic stakeholders (clinician researchers; *n=2*). This sample size was based on recommendations for traditional co-design practices [17], and also on the practicalities of “seeing” everyone at once in the online sessions. The choice was made to have higher numbers of end-users (parents) as part of efforts to recognise and address potential power imbalances that could occur within the group.

Parents brought their experience of parenting an extremely preterm child, and their experience of accessing and participating in recreational physical activities with their preschool-aged child in Victoria, Australia. The two circus coaches brought their experience of teaching circus to preschool-aged children and of managing preschool circus programs in two diverse locations in Australia, one a small independent company in a rural town and the other a major capital city in one of the largest circus schools in the country. Furthermore, one coach had a background in trauma-informed education, and the other had training in occupational therapy. One clinician was a paediatric psychologist working in a hospital setting and a not-for-profit Early Childhood Approach National Disability Insurance Scheme setting. The second clinician was a physiotherapist with a special interest in paediatrics working in a private practice setting. The academic stakeholders were both researchers and physiotherapists with backgrounds in teaching recreational physical activity (circus: F.C., and dance: C.M.) to young children. One researcher had some prior experience in co-design (C.M.), and both had experience in qualitative and mixed-methods research. F.C. was the project lead and workshop facilitator with C.M. providing facilitation support during workshops.

Informed consent to participate in the project was sought in writing, with consent for interviews reconfirmed verbally by the interviewer. End-users and service-based stakeholders were reimbursed AUD$50 per workshop, one academic stakeholder (F.C.) provided their time in-kind as part of their Doctor of Philosophy (PhD) degree, and the other academic stakeholder (C.M.) was reimbursed at AUD$50 per hour.

### Data collection

#### Part 1: Co-designing the rehabilitation intervention

The P-POD process was conducted from June to September 2021, with team members (*n=*10) participating in diverse ways (described in Table 1), but primarily through eight 90-minute online workshops. After an initial five workshops, the group voted to continue for another three, for a total of eight sessions. The additional three workshops were indicated to finalise key intervention details. As these added workshops required additional time, team members were given the option to continue participating, with seven team members opting to continue. Detailed agendas for each workshop can be found in Padlet (https://padlet.com/free_c/session_notes).

#### Part 2: The evaluation of P-POD

The few studies reporting comprehensive evaluations of co-design and stakeholder involvement processes have used a variety of data collection methods, including semi-structured interviews [5,45], focus groups [37] and email feedback [46]. For evaluation of P-POD, anonymous online surveys and semi-structured interviews were selected. The final survey and interview question guides can be found in Appendix 1, and were developed based on recommendations from the literature related to evaluating co-design [5,8,10,17,22,37,45,47].

Surveys were utilised to evaluate the adherence of each P-POD session to the guiding principles of the process, satisfaction with the session, and suggestions for quality improvement for the subsequent sessions. The anonymity of the surveys was felt to be important to reduce social desirability bias, allowing participants to respond more freely [48]. Having 5-point Likert scales to rate adherence to principles, satisfaction, and engagement with the session made the surveys quick to complete (less than five minutes), reducing the burden on team members, while still capturing useful data. Surveys were administered via REDCap (Research Electronic Data Capture version 12.5.16 hosted at Murdoch Children’s Research Institute, Melbourne, Australia [49,50]) to all attending team members at the end of each online workshop (June to September 2021).

Optional semi-structured interviews (offered via phone or video-conferencing) were selected to explore participants’ experiences of the online co-design process. A phenomenological approach informed the development of the interview questions, including positive aspects and challenges, goal achievement and perceived learning and impact [51]. Interviews were conducted by K.C. after the conclusion of the P-POD process (September-December 2021). K.C. had not been involved in P-POD but had experience in qualitative research and interviewing.

### Data analysis

#### Part 1: Co-designing the rehabilitation intervention

The resulting co-designed intervention was mapped into a TIDieR checklist format during the research co-design process (see Appendix 2) as a guide to ensure complete reporting of the intervention design [52]. This data has been summarised and presented in the results section below.

#### Part 2: The evaluation of P-POD

Quantitative survey data were downloaded from REDCap and imported into Microsoft Excel for Mac (2023; version 16.71). Descriptive statistics were performed by F.C. under the guidance of a statistician. Evaluation interviews were audio-recorded and transcribed verbatim by OutScribe (an external transcription company). All identifying information except for stakeholder type was removed prior to the reflexive thematic analysis of the qualitative interview data being undertaken in six phases as outlined by Braun and Clarke [53], described in detail below. Member checking was undertaken by interviewees reviewing the themes and being asked if the results resonated with their experience and if there was anything they would like to add or change. No changes were requested to themes, and participants reported that themes resonated with their experiences.

##### Step 1: Dataset familiarisation

All analysts familiarised themselves with the entire dataset by listening to the audio recordings and/or reading the transcripts. F.C. made notes of initial insights that arose during this stage (both throughout the data and of the dataset as a whole). C.M. and K.C. independently followed a similar approach for a subset of the data (two interviews each), and for the remaining data, they each noted any key differences or similarities to enhance understanding of the full dataset.

##### Step 2: Data coding

An inductive approach was used to facilitate an empathic, experiential approach to the analysis. Interesting or meaningful segments of data that appeared to answer the research question, were labelled with one or two words that captured a key concept or idea. F.C. coded all interviews, while C.M. and K.C. independently coded two interviews each. Qualitative data analysis software NVivo v12 [54] was used to organise and manage the data through the coding phase.

##### Step 3: Initial theme generation

All three analysts met to review the codes generated in Step 2. Collaboratively, they grouped codes that appeared to share a core concept and illuminated an aspect of the research phenomenon. This resulted in five candidate themes, with associated coded data.

##### Step 4: Theme development and review

F.C. synthesised the associated data for each theme and prepared a draft for review. The analysts met a second time to assess the fit of the candidate themes to the overall analysis by checking the relevance of the candidate themes to both the associated coded extracts and the full dataset. They also considered the scope of each theme and clearly articulated what would be included, and not included under each, resulting in four final themes.

##### Step 5: Theme refining, defining and naming

F.C. then prepared another draft, and all three analysts met a third time to review and further refine the themes, their descriptions and finalise theme names.

##### Step 6: Writing up

F.C. prepared the draft manuscript with all authors providing feedback and revisions.

#### Reflexive Statement

This statement aims to inform the reader as to the actions taken to sustain a reflective practice and describe influences on the interpretation of the qualitative data. Appendix 3 provides a more detailed reflexive statement from the project lead (F.C.). Reflexivity was sustained throughout the design and conduct of this research by reflecting during fortnightly meetings with the core research team (F.C., A.S., R.T. and K.S), and via meetings and email contact between the co-analysts (F.C., C.M. and K.C) during the qualitative analysis and write up stage. Although having more than one analyst is not considered necessary in reflexive thematic analysis, due to the involvement of F.C. in all aspects of the co-design and evaluation, a collaborative approach was taken to ensure that the analysis remained open to different perspectives and to enhance reflexive processes [53]. All three analysts have experience in qualitative research methods, with C.M. and K.C. having specific experience in Braun and Clarke’s reflexive thematic analysis [53].

All three analysts are physiotherapists with backgrounds in coaching recreational physical activity for children. They place significant value on inclusive opportunities for physical activity, with both F.C. and K.C. having undertaken PhD degrees on this topic. Furthermore, F.C.’s two-decade experience teaching circus activities meant they had preconceived notions of “what works” and worked consciously through the research co-design workshops to not utilise their experience as a coach to influence the design, and to prioritise the perspectives of the other circus coaches present. C.M. and K.C. also have prior experience in research co-design. Along with all three analysts’ passion for genuine inclusion in co-design, during the analysis attention was paid to negative cases and suggestions for improvements to P-POD were incorporated into the thematic findings.

## Results

### Part 1: Co-designing the rehabilitation intervention

The resulting co-designed intervention was titled by the co-design team as “CirqAll: Preschool Circus for Premmies” and was a three-component intervention (see Figure 3). The aim of CirqAll is to meaningfully include preschool-aged children born preterm in circus-based recreational physical activity, while enhancing social and physical development. A TIDieR checklist [52] describing the full intervention design developed by the co-design team is provided in Appendix 2.

**Figure 3:**
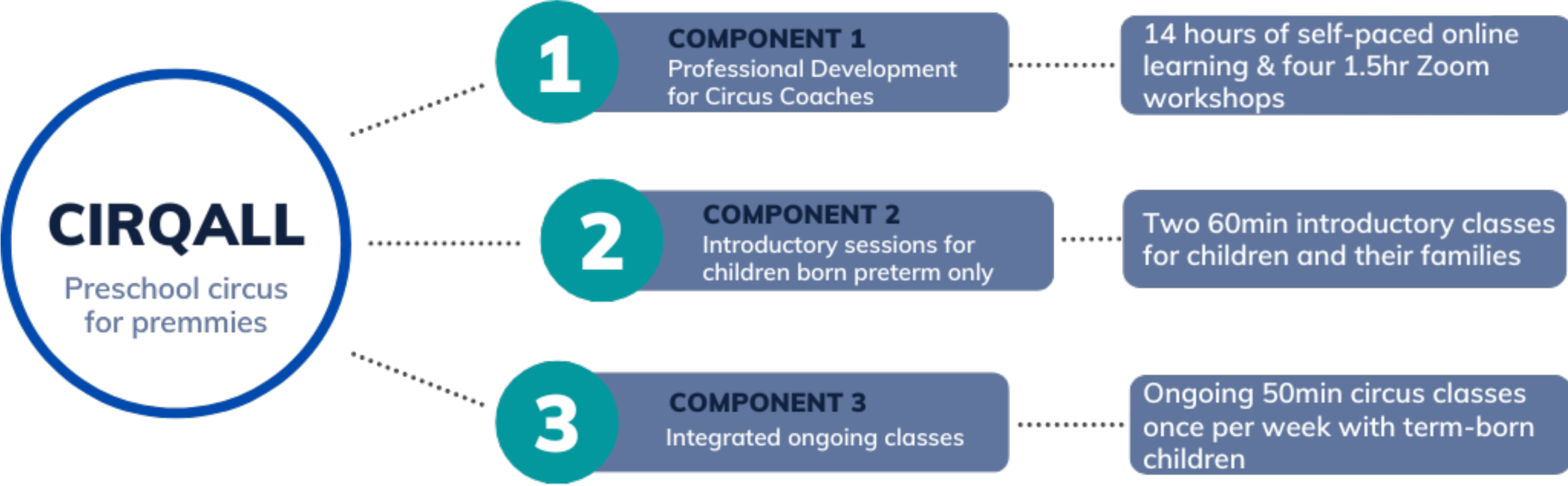
Components of CirqAll, the co-designed intervention. Figure 3 Alt Text: A heading titled CirqAll: preschool circus for premmies, with three branches showing the three components of the intervention.

In Component 1, circus coaches complete professional development to extend their existing capacity for working with diverse needs into a preterm-specific context. Increasing coaches’ confidence and expertise in enhancing participation and developmental outcomes for children born preterm was identified as a key need in a prior study undertaken by the authors [44]. The content of this professional development was developed from the analysis of data from this prior study, and further refined by the co-design team. In Component 2, children born preterm (and their families) attend two 60-minute introductory circus classes with other children born preterm to become familiar with the coach, the class environment, and foundational circus activities. Furthermore, during these sessions, time is allocated for coaches and parents to discuss individual strategies to support their child for optimal participation during Component 3. Component 3 is a weekly 40-50min circus class in a continuous term-based program, integrated with term-born peers.

### Part 2: The evaluation of P-POD

As illustrated in Table 2, forty-six survey responses were collected from the co-design team over the eight workshops. The first author (F.C; an academic stakeholder (ResearcherA) and project lead) did not complete any surveys, as they felt too tightly bound to the evaluation process. Eight of the ten team members consented to the optional evaluation interview (parents; *n=*3, coaches; *n=*1, clinicians; *n=*2, academic stakeholders; *n=*2). Interviews had a mean duration of 37 minutes (range 20-50 minutes).

**Table 2:** Participant evaluation data

| ***Co-design***<br>***Team Members*** | **End users**<br>**(parents *n*=4)** | **Service-based**<br>**stakeholders**<br>**(clinicians *n*=2;**<br>**coaches *n*=2)** | **Academic**<br>**stakeholders**<br>**(researchers**<br>**(*n*=2)** | **Not identified<sup>1</sup>** |
| --- | --- | --- | --- | --- |
| *Survey responses*<br>*(n=46)* | 18 | 19 | 7 | 2 |
| *Interviews (n=7)* | 3 | 3 | 2 |  |

Table notes:
<sup>1</sup> Team members who did not indicate their stakeholder type in the survey responses

Satisfaction and engagement with the workshops were high, with 45/46 responses indicating that team members were all, or mostly engaged and satisfied with every workshop (Figure 4). Furthermore, 44/46 responses indicated that team members felt they were gaining new skills or knowledge that was useful during the sessions.

**Figure 4:**
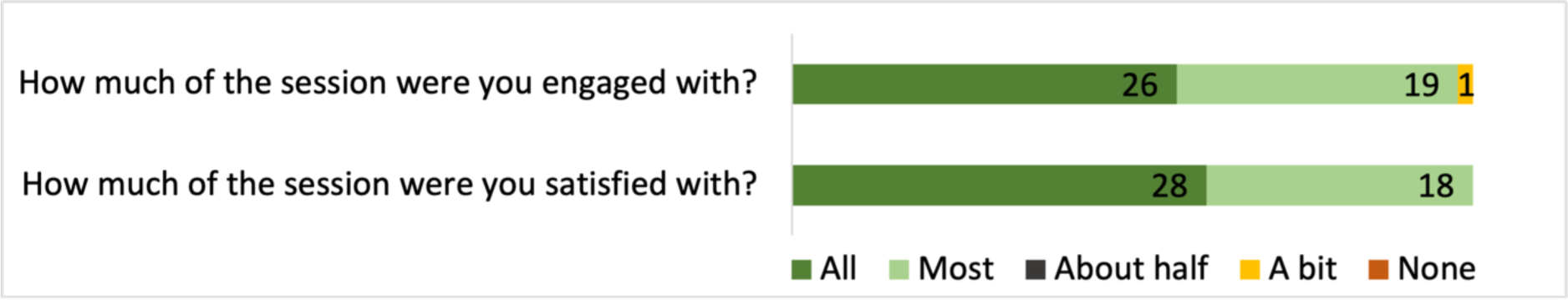
Anonymous survey responses related to participant engagement and satisfaction with the sessions. Figure 4 Alt Text: A bar chart showing that 45 out of 46 responses indicated all or mostly satisfied and engaged with workshops.

Adherence of workshops to guiding principles was high, with more than 80% of responses indicating ’agree’ or ’strongly agree’ to each statement (Figure 5). In the first workshop, one team member selected ’strongly disagree’ to all statements with no quality improvement suggestions provided. However, when asked how satisfied and engaged they were in the session, they selected ’a lot’ for both responses and responded ’yes’ to the question asking whether they felt they were gaining new skills or knowledge that was useful. Another team member disagreed with the statement “I felt like the session achieved its objectives” for one workshop.

**Figure 5:**
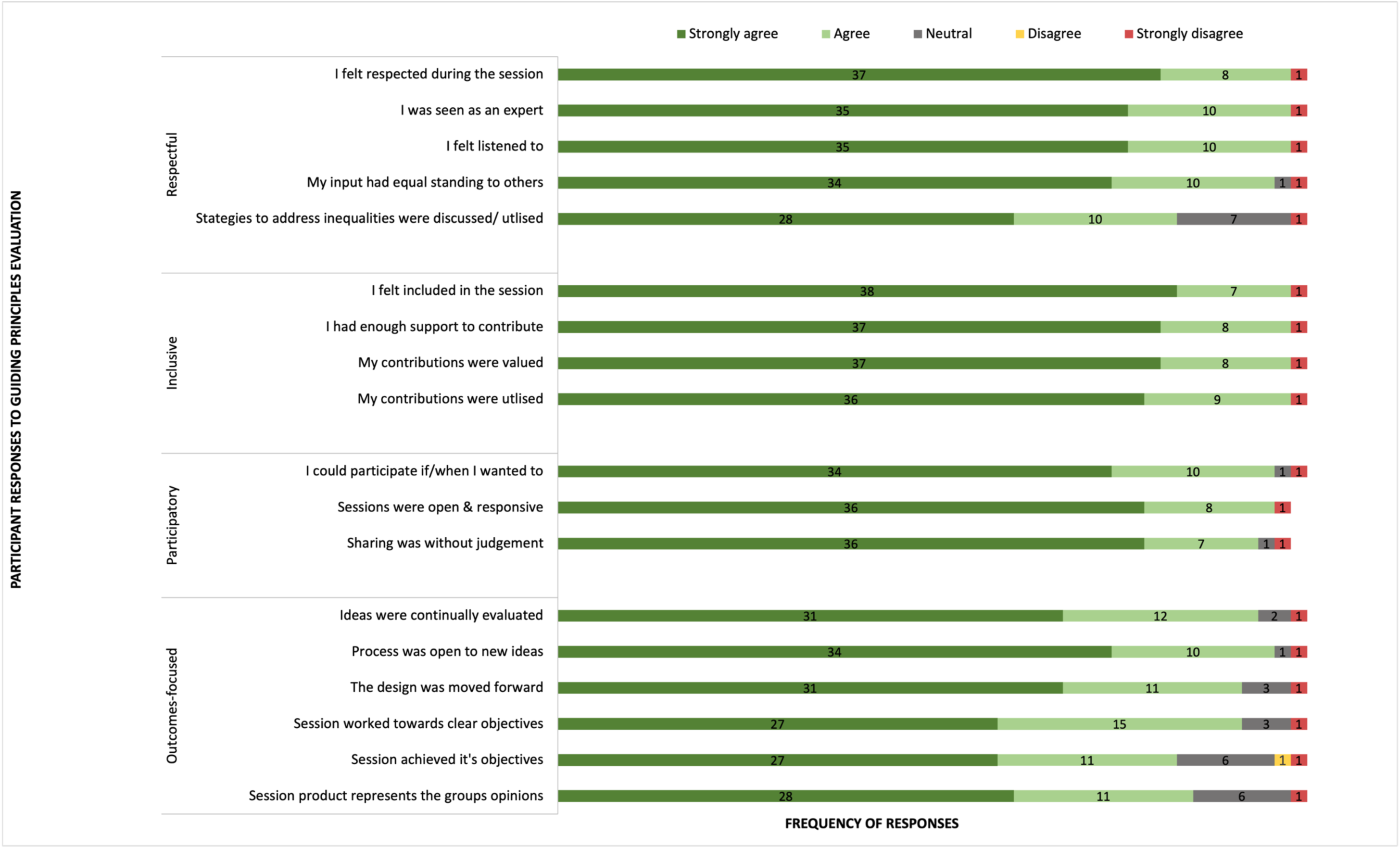
Anonymous survey responses related to the adherence of workshops to guiding principles. Figure 5 Alt Text: A bar chart listing statements relating to adherence of workshops to co-design principles.

Four themes were developed from the interview data which explored team members’ experience of the P-POD process:

1. Co-curating the online culture,
2. Difference without discord: valuing diverse perspectives and “making room for healthy debate”,
3. “Everyone is equal” but…power in P-POD,
4. Defining success in P-POD.

### Theme 1: Co-curating the online culture

Team members described P-POD as having a *“really nice culture”* (CoachA), which was felt to be genuinely collaborative, inclusive, and *“very respectful”* (ParentB, CoachA, ClinicianB, ResearcherB). This theme describes participants’ reflections on particular elements that curated this environment, including the work done prior to commencing P-POD and the initial P-POD sessions. Tools used to develop this culture included: building relationships, encouraging participation, assembling an invested team with the required expertise, developing guidelines for the group, and the use of online platforms. The term ‘co-curating’ is used in the theme name to describe participants’ reflections on how their contributions also helped to shape the group culture.

The use of a video conferencing platform as the online co-design environment was viewed by participants as convenient and enabled an inter-state team to participate. One participant commented *“I’m all about Zoom. I never want to go to a face-to-face meeting ever again. I love this”* (ClinicianB). Other participants felt that a hybrid delivery may have enabled a more *“natural”* (ParentC) conversational manner which may have furthered relationship building. Two participants reflected that the positive *“flip side”* (CoachA) of this *“artificial environment”* (ParentC) was that participants could have breaks when needed without drawing attention to themselves and that sessions were very productive: *“We get into it and we get working”* (CoachA).

### Sub-theme: “All the background work behind it”: setting the scene for collaboration

Distinct and purposeful actions were taken in the months leading up to the co-design process to curate a respectful, collaborative online environment. One key action was undertaking a prior mixed-methods study to understand the needs and preferences of stakeholders relating to a circus intervention for the preterm cohort (44). This initial study enabled key data on the clinical problem to be collected, informed the vision and goal for the co-design process, and facilitated the sampling of invested team members. Participants credited this study as resulting in a clear vision and goal *“to frame the co-design process”* (CoachA) as well as providing *“rigorous data that we could draw on to inform some of our discussions and decisions”* (ResearcherB).

Relationships within the team were described as integral to the group culture. Participants identified that meeting with the project lead (ResearcherA) and other participants in a prior focus group helped to quickly build relationships with the co-design team. They described how these prior interactions meant that they also felt clear about the vision of the project from the outset.

> *It was nice to chat to [ResearcherA] before as well, so when we came to Zoom it was familiar too […] it was nice to see some of the parents too from the focus group. Because you could feel that familiar[ity]* (ParentB).

The use of breakout rooms via the online platform was also credited with further developing relationships between the team *“because we could just chat […] it made you closer and more understanding of different people’s backgrounds which made the group gel more”* (ParentB).

Another key aspect of the *“background work”* was identifying and bringing together a team with a shared commitment to the co-design process. This was managed through purposive sampling of co-design team members to bring together an invested and enthusiastic group with the required expertise (both professional and lived experience). Participants commented on these attributes influencing the culture through the group’s enthusiasm and commitment to the process.

> *This level of engagement and enthusiasm, and a willingness to invest time in the project. […] I think they’re really powerful attributes to have in a research team, all working collaboratively. And I think the attitudes that people came with helped to set up some of that respectful listening, and then they were reinforced by some of the initial sessions* (ResearcherB).

### Sub-theme: Early content crucial for “actively creating that environment”

Many participants reported that the first couple of P-POD workshops were instrumental in setting up the culture. In workshop 1 an exercise was conducted using online collaborative software whereby the team constructed clear expectations to govern conduct during meetings. This was felt to contribute to the early development of the participatory, collaborative, and respectful culture, and these guidelines were referred to at each meeting to remind participants of the team’s shared values and expectations. *“At the beginning everyone came up with sort of values [..] like a guideline that everyone sort of contributed to. I think that was helpful and that was something that was always visible”* (ClinicianB).

Learning about the co-design process itself was also reflected on by participants as instrumental in setting the culture and expectations. *“[ResearcherA] really took the time to talk about how this process works and how the design team is programmed to operate, I think that was helpful actually”* (ParentA). Specifically, introducing the expectation early of active participation throughout the process influenced the environment strongly. *“She really, from the get-go, went ’Come on, participate’”* (ClinicianA). Introductions between the team members to get *“an idea of everyone’s backgrounds […] so you can sort of understand where everyone’s points of views are coming from”* (ClinicianB) also contributed to the environment of respect as emphasis was placed on lived and professional experiences being of equal importance.

Team members also reflected on their work in further shaping that *“very collaborative environment”* (CoachA).

> *…that environment that we put together, I totally give [ResearcherA] credit for helping create that, because I know it has to come from there. The groundwork she did in week 1 and 2 definitely helped us get to that space as a group, that we were really positive and collaborative, and really those positive expectations of everybody was good* (CoachA).

One participant noted a distinct journey in the team culture moving forward: *“[at] the start it was all like strangers or tiptoeing, and then as it got in, we all sort of became like a group, just like a friendship group, catching up every week and continuing as a team with a shared goal”* (ParentB).

In summary, participants’ reflections on the unique benefits and challenges of the online environment and how the group culture was curated included: the use of the online platform, attributes of the assembled team, the background work done prior, and the content of the initial workshops.

### Theme 2: Difference without discord: valuing diverse perspectives and making “room for healthy debate”

This theme describes how participants valued the diverse team members and their contributions and spoke of how they enriched and challenged each other during workshops. This experience was potentially enabled by, and related to, the respectful culture described in theme 1. All team members described feeling that their contributions were valued by the group throughout the process. One participant even described how they had developed confidence in their stakeholder expertise through participating in P-POD.

> *“You kind of just do what you do as a parent and you don’t realise that it might be helpful to share that with other people. So this co-design process gave me some confidence there […] that’s what this co-design process made me see is that, what we do can be quite valuable and that it might be helpful to other families”* (ParentA).

All participants described valuing the diverse backgrounds and perspectives of the team members and *“the exposure to all the other people, the other stakeholders that were engaged with this, their understanding and views on all the different little specifics”* (CoachA). The team’s *“really big mix of backgrounds”* (ClinicianB) was mentioned by all participants as having a significant impact on their experience in P-POD. One participant expressed that the wealth and diversity of experience in the team relieved some feelings of anxiety about the complexity of the process and the breadth of expertise (both lived and professional) required to achieve the project’s goal: *“[…] it’s not meant to rely on one person with expert knowledge. It’s about drawing on everyone’s experience and expertise”* (ResearcherB).

Many participants mentioned the parents’ contributions and perspectives as being *“the most valuable part of it”* (ClinicianB), as their insights into potential solutions based on their experiences of engaging their child in recreational physical activities were honest and thoughtful. However, one participant noted the differing levels of vulnerability required when sharing lived experience (end-user stakeholders) versus professional experience (academic and service-based stakeholders).

Parents spoke of their appreciation of having other parents in the team, and how the shared (but diverse) experiences voiced by other members of their stakeholder group were felt to validate their input. *“I could relate to those sorts of experiences and not feel it was just me or why is this because of or why is this just my son or whatever, that sort of mindset. I almost felt validated”* (ParentC).

Even within a diverse group of parents where *“not everybody’s experience with a premmie was the same”* (ParentC), a couple of participants highlighted that a risk of co-design is that the resulting design may be *“a little bit limited by [participants] own experience of what had and hadn’t worked”* (ClinicianA). These team members did not feel this had negatively impacted the process, but “*it just made me wonder if we were missing maybe some of the wealth of experience that was out there”* (ClinicianA).

> *I don’t know if it’s a negative but there were times where some parents were not necessarily focused on the bigger picture, but on what they needed for their child. […] And maybe that was part of what they were supposed to be doing, you know just really focussing on… they only know what they know for their own child right? It’s not really a negative, it was just something that I noticed.* (ParentA)

The diversity in backgrounds and experiences of the team meant diversity in perspectives and opinions too, and for some team members, the differing opinions shared in the process were unexpected: *“I probably wasn’t expecting the range of different ideas and topics we were talking about and some people think so different to what I was thinking which is good”* (ParentB). All participants spoke of these diverse opinions positively and felt they *“enriched the experienc*e” (ResearcherB). There was a strong sense of inclusion of each person’s perspectives throughout the process. *“There might have been healthy disagreement, but there was always this validation of the thoughts […] everyone’s thoughts and opinions were considered. Nobody was discounted or excluded from being able to give an opinion”* (ParentA). This sense of difference without discord and non-judgemental sharing of opinions was articulated by all participants.

> *…there wasn’t any arguments or whatever. But it was like oh yeah, I see how you see that, I see how you feel but I feel like this. And we were all very respectful to how everyone thought, what ideas they had […] you didn’t feel like you were judged if you had a different opinion, not that we really had that vibe anyway.* (ParentB)

Participants appreciated the *“room for healthy debate”* (ParentA) where people could express their opinions and perspectives and be challenged without judgement by another team member’s differing perspective. Participants described how these challenges to ideas and assumptions were positive and meaningful enough to prompt them to reconsider their stance, whether that resulted in re-affirming their perspective or led to a change of opinion.

> *I had an idea of what I wanted, but once we batted it round the group, I was like, “Yeah, no, I’m wrong. We should do it this way. That’s going to be way better.” Repetitively. And I think that happened to pretty much everyone at some stage, we were like, “We should work this way”, and then a couple of minutes later, “No! It’s changed our mind. Yeah, that was the wrong expectation. I had an assumption there. I needed to change it.” I don’t see that happening as much in traditional groups as it happened in this one. It was cool.* (CoachA)

The journey through the *“healthy debate”* was viewed positively by participants, and *“always, always, consistently approached really collaboratively and respectfully”* (CoachA). One tool that participants felt very strongly enabled this collaborative and respectful coming together, was the Consensus Decision-Making framework (55), which was introduced in workshop 3 and used regularly for the remainder of the process.

> *Everyone was okay once they’d had their say, people were happy to move in one direction eventually, so there was consensus, and we overcame it through that decision-making model I think. There was never anyone shouting or getting worked up and I think that’s a credit to the team and to the framework that was put in place.* (ParentA)

Participants described how this framework’s approach aligned with the values of P-POD, particularly in supporting a respectful collaborative way to move the design forward. *“I liked the process in that it never really came to a head, it was just a process of continuing to discuss our viewpoints and then coming to [consensus]… ultimately, we weren’t forced to choose a way forward”* (ClinicianA).

The clear goal of the process and the session-based agenda also seemed to play a role in helping the group keep the design progressing, even when participants had strong opinions on aspects of the design that took time to work through. Participants also commented that both the process and the end design output were able to cater for multiple perspectives without significant compromise: *“it ended up in a really good place in the design process, because we were able to cater for both and find that nice middle ground for everyone”* (CoachA).

### Theme 3: “Everyone is equal” but…: power in P-POD

This theme describes participants’ reflections on the power dynamic and the sharing of power in P-POD. They described having power in decision-making and design direction, however, they also described ResearcherA as having a distinct facilitation and coordination role that was essential to the process. Actions taken by the academic stakeholders to address power imbalances by sharing tasks and roles were appreciated but not always taken up by participants.

### Sub-theme: Stepping forward and developing agency

Some participants described their surprise at the power dynamic in P-POD, particularly how ResearcherA as the project lead *“stood back”* (ClinicianA), held space, and gave support for the team members to ’step forward’ into a collaborative process. Participants expressed appreciation for the amount of control the group had over the process, *“the power to direct where it’s going”* (CoachA) and felt this was set up early in the process and facilitated throughout. They reflected that P-POD was unlike other experiences of more *“traditional”* (CoachA) meeting structures where “*you expect that someone’s going to chair it and lead it and that you’ll be silent until you’re asked for input, whereas right from the start, [ResearcherA] explained that that wasn’t how this would be. [..] she made an effort to go she wasn’t the one leading it. This was the group leading this discussion”* (ClinicianA). As a result, participants often described a sense of ownership of the design output. *“I have a genuine sense of involvement and inclusion in what we’ve created and a genuine sense of buy-in to the success or the failure of the project”* (ParentA).

Many participants also described the Consensus Decision-Making framework’s role in addressing potential power imbalances and providing agency to team members in crucial design choices. *“It wasn’t someone making one decision, it wasn’t like [ResearcherA] going okay well the vibe of the group is this, we’re going to go this way. It was really fair I think, fair for everyone”* (ParentB). In reflecting on their experiences in other co-design processes, ResearcherB commented that in times of key decision-making typically they had experienced a ’stepping in’ *“by the lead researcher making a key decision”*, whereas participants in this process reported:

> *It wasn’t like, I’ll give my opinion but you’re all going to make whatever decision you want […] this was a genuine respect and inclusion of everybody’s thoughts and opinions that was going to drive the outcome of the project, which again, just blew me away that that could be successful. And that that could work* (ParentA).

Participants also described an appreciation of their agency regarding their individual input into the process, being able to choose when and how their experience and perspectives were contributed. Participants reported that they did not feel compelled to contribute, but if they had something important to say, they could say it through a variety of methods.

> *You felt like if you had something to say you could say it […] if you don’t want to say it on the screen, email [ResearcherA] or if you don’t want to, you could put it in that question box at the end of each session. So it was very much what you’re comfortable with.* (ParentB)

Participants mentioned ResearcherA’s active role in *“making sure that we checked in with a couple of people who weren’t talking as much”* (ClinicianA), and that small group discussions and activities (via breakout rooms in the online platform) assisted in making *“sure those voices were heard”* (ClinicianA). Participants also appreciated an active seeking out of each person’s contribution if unable to attend the group sessions. *“If you couldn’t be on a meeting for whatever reason, [ResearcherA] gave her time to go and search out people’s thoughts outside of that meeting”* (ParentA). For one participant who chose to contribute primarily via email *“it felt that [ResearcherA] absolutely made an effort to contact me and get my opinion. Whilst it’s not as effective as being there in person, at least I was able to provide some help or provide my experience or my thoughts in other ways. So I think that could absolutely help with the project”* (ParentC).

### Sub-theme: Power in, and sharing of, roles in P-POD

Even though participants reported agency in contributions, design discussions and decision-making, there was a clear distinction between different levels of power in the roles of session facilitator and project lead. As project lead, some power was retained by ResearcherA as they *“designed that process. So that’s where she really gets the power to direct this”* (CoachA). As ResearcherA being *“the only one with that full vision”* (CoachA), some participants described feeling like they lacked a sense of *“the bigger picture”* (CoachA) and expressed interest in seeing *“a bit more of a map of where each co-design meeting was going to go […] I don’t think I really had a good picture of that from week to week, but it didn’t matter”* (ParentA).

The role of session facilitator was commented on by many participants as being essential to progressing forward in the design process by keeping the discussions on track and valuing the time of the team members.

> *Although it’s a group discussion and everyone is equal, I feel like you actually do need someone whose role and responsibility is the moving on or the redirecting [..] I think you still do need that one person sort of guiding or help redirect as needed* (ClinicianB).

Offers to share this role with other members of the team were made by ResearcherA in an attempt to address power imbalances, with participants appreciating these attempts but expressing mixed perspectives on how successful this role-sharing was in practice.

> *…it wasn’t always [ResearcherA leading the session]. Sometimes she’d ask someone else to run the session, like be the session person.* (ClinicianB)
>
> *She tried to hand over that lead as well. No-one picked it up. Everyone was like, “Oh my gosh, no-one’s going over there”.* (CoachA).

Participants reflected that the action of creating opportunities for role-sharing allowed *“the rest of us to step forward a bit more than we would have, I think”* (CoachA). However, one participant questioned if enough support had been provided to team members to take up those roles, or whether some improvements could be made to this power-sharing strategy.

> *If the reason was that they felt that they had genuine choice and didn’t want to, that’s great. I would say then we achieved what we set out to achieve, offering the opportunity and giving people choice. But if there were other barriers, like someone didn’t feel confident or they wanted more support or training to step into the role, outside of an online session, or they wanted a more clearly defined role, so they really felt confident around when to step in during the session, those would be aspects that would, to me, be a marker of maybe didn’t quite get that part of it right, that we were offering opportunity but not giving people enough support to really take up that opportunity* (ResearcherB).

There were also organisational and administration tasks done between sessions, with participants noticing and valuing *“how much work was put in behind the scenes as well”* (ParentB). *“I assumed that [ResearcherA] had to do more outside of the co-design team. Like otherwise it wouldn’t have all come together the way that it did, but somebody had to, and if [ResearcherA] could, that was great”* (ParentA). Participants felt that the organisation around the process *“was really critical for enabling me to feel like I could contribute well to the sessions […] If it hadn’t been that organised, I suspect it would have been really hard for me to contribute in a meaningful way”* (ResearcherB). Participants expressed appreciation for this additional work, and although many team members participated between sessions by contributing to the Padlet, or reading minutes, some participants reflected that taking on the administration tasks wasn’t possible as they were already at capacity and *“time-poor”* (ClinicianB).

In summary, participants acknowledged attempts to address power imbalances in P-POD, and overall, described a sense of agency throughout the process.

### Theme 4: Defining success in P-POD

This theme describes how participants defined and experienced success in P-POD, both regarding the pre-defined goal of designing an intervention, but also the satisfaction of individual motivations to participate, and key takeaways that participants intend to integrate into their personal and professional lives beyond the project. *“I think it definitely helped me grow as a person definitely, which is kind of weird to say in like eight sessions. But yeah, it was quite enlightening […] it’s opening my world to different things”* (ParentB).

### Sub-theme: The project goal: “big tick” but “next stage” essential

All participants felt that the co-design process had been successful in its goal ’to co-design an intervention using circus activities that improves physical activity participation for pre-schoolers born preterm, using the information collected from parents, health professionals and circus coaches, and the combined expertise in the Co-design Team’. *“I like to think that we’ve got a big tick against that goal. Yes”* (ParentA). Some participants described wanting to continue for longer in the design process but questioned how to define the end point “*eventually you have to cut it off somewhere and then use what you’ve got otherwise it will just keep going and going”* (ParentB). They also reflected on the tension between further work in P-POD, and their actual capacity to commit more time to the process, as well as whether more time would have resulted in a better project outcome.

> *Maybe more time? Probably not, actually […] we could probably continue forever talking about it, because we’re all pretty into it and engaged. But I think after the 8 weeks, yeah, there’s something there that will absolutely tick those boxes, absolutely improve physical activity and participation in pre-schoolers, yeah* (CoachA).

There was also a strong sense from many participants that the implementation of the designed intervention was a critical part of being able to definitively say the project was a success. *“Sometimes these things can sound good on paper, but whether it actually works in practice will be another question”* (ResearcherB). Participants were invested in this next phase and expressed sincere hope that implementation is achieved. *“I’ll be really disappointed if it doesn’t because I think we’ve all worked so hard on it”* (ParentB).

### Sub-theme: Giving back, paying forward: the cycle of reciprocity

All end-user stakeholders reflected on feeling *“content”* (ParentC) that they had successfully satisfied their motivation for participating in the process, which for all parents was related to *“wanting to give back”* (ParentA). Many parents described having benefitted from the results of prior research regarding optimising health and wellbeing for children born preterm and wanting to *“pass it along”* to *“help a premmie now or a premmie in five years*“ (ParentB). Further, they appreciated the joint motivation present in their stakeholder group of *“very strong parents*“ (ParentC).

> *I felt good that I was able to be a part of a team of very strong parents who have been through a similar situation that I went through at the time, and we could all come together and help out in any way that we could […] I’m really inspired by strong women and for me, I guess having some really strong people within that team of mums of children born preterm, it really inspires me and it really did inspire me listening to some of the stories. I take that away as a way to make me stronger and I think that again, maybe that it’s another advantage that I had from being a part of this group… you see other mums also talk about that and you’re listening to their experiences and it’s just really inspiring* (ParentC).

### Sub-theme: Impact beyond the project

All participants described an appreciation of gaining transferable skills and knowledge that they were intending to integrate into their professional and/ or personal lives. The clinicians reported that learning more about circus has prompted them to consider referring families to circus programs and to *“suggest them to families over and above some of the other more traditional sports programmes […] what I’ve heard about circus already, I think it’s probably more adaptable and more welcoming […] I think I’d be more keen on circus than other group sports programmes for kids”* (ClinicianA). Both health professionals also reflected that the process had re-affirmed for them the importance *“of what’s going on for parents, what their perspective is and how important that is to inclusion”* (ClinicianA) and how programs can be modified to ameliorate barriers for parents and *“reduce that task or burden ongoing as well”* (ClinicianB).

Many participants spoke of utilising the Consensus Decision-Making framework moving forward, both in their professional and personal lives. They spoke of its value in regard to honouring diverse perspectives, but also as disrupting the traditional power structures in key decision-making processes.

> *I think that whole concept of decision-making in a very different way has filtered through for me. Yeah, I’ve been trying to be more, what’s the word? Collaborative in the way I make decisions with my older children, one’s a tween and one’s a teenager. At work, I’ve been really trying to back off from being the decision-maker and collaborate with everybody in a different way. The validation and the respect of other people’s opinions is something that you don’t see very often […] I don’t see it that much in everyday life and I really like it. I can’t say I do it all the time but I’m trying to use that in my life* (ParentA).

Many participants also reflected on their learnings of co-design as being a strong takeaway, and some described how they would integrate co-design into their future professional lives.

> *I’m a project manager myself, so just having another view of how you can work with stakeholders and engage people in a really collaborative way that minimises that power imbalance between possible relationships, is always a good, practical experience for me* (CoachA).
>
> *I was just talking about it with a friend before, I would really like to go somewhere with this somehow in my life. I don’t know where, and I don’t know what that looks like, and I’m kind of happy for it to be organic, not to drive it* (ParentA).
>
> ResearcherB as an academic stakeholder spoke of their reflections prompted by this process around what authentic co-design is, or isn’t, and how they would share their experiences and knowledge in the future to respectfully educate other researchers wanting to engage in research co-design.
>
> *It’s also just reminded me that there’s a lot of tokenistic consultation that goes on, that’s masked as co-design […] How might I use some of these learnings from this co-design project to help share my knowledge and experience of what co-design is and how it differs from consultation, and how, as a research community in particular, I think we really need to make sure, when we’re saying we’re doing co-design, we really are doing co-design. I think that’s important from an ethical perspective, actually* (ResearcherB)

## Discussion

This study found that a novel online approach to research co-design (P-POD) can produce a co-designed rehabilitation intervention (CirqAll: preschool circus for premmies) that addresses stakeholder needs identified in the authors’ prior work [44]. Furthermore, quantitative survey findings demonstrated that stakeholders participating in the P-POD process found it satisfying and engaging and felt that it adhered to pre-defined guiding principles. Four themes developed from the qualitative data described participants’ experience of P-POD, such as the respectful online culture, the importance of differing perspectives and room for debate, how success was perceived in multiple ways, and how power was perceived as being shared but not equal.

There is a scarcity of literature evaluating online co-design approaches. Furthermore, substantial differences in online co-design processes and levels of stakeholder involvement make a comparison between findings difficult. For example, one co-design study described participants completing a one-off feedback task to improve a patient-facing area of a hospital, with no interaction between participants, or with a facilitator [56]. In another study participants also only attended one session (there were two workshops in total held with different stakeholder groups), however, the research team utilised activities prior to the workshop to familiarise participants with the objective and key content for the co-design [25]. The authors reported this pre-work allowed an accelerated start to the co-design, which was similar to the benefits that P-POD participants reported from their involvement in the prior information-gathering study.

Fails et al. [24] conducted several online co-design projects with children and found trouble-shooting technology difficulties to be a major theme across their projects which were not identified as a challenge in P-POD, perhaps due to the age of the team members. Although power was raised as a major theme in both P-POD and Fails et al.’s work, in the latter, the authors found it harder to disrupt the child-adult dynamic in an online environment, due to children requiring adult assistance to work through technical issues. Fails et al. [24] also reported that the in-person power dynamic was more equitable, as children could have more agency over their participation, choosing when and how to participate as well as being able to take breaks more easily and with less disruption to the group. In contrast, P-POD participants highlighted their agency in contributions and the ease of taking breaks in an online setting as key experiences of the process.

In light of this limited evidence base, the objectives of this paper were to describe and evaluate a novel online research co-design approach (P-POD) to understand if it is an authentic way to transition research co-design into an online environment. To meet this objective, the mixed P-POD evaluation data is mapped back to the guiding principles as identified from the literature. The mixed data is then interpreted and contextualised within contemporary literature on in-person co-design approaches to provide clear recommendations to researchers to capitalise on the accessibility and convenience of online approaches while upholding the authenticity of co-design principles.

P-POD defines the principle of *being respectful* as an equitable partnership that sees all participants as experts, aims to recognise and reduce the effects of power imbalances, and engages in shared decision-making to produce a collaborative design. P-POD’s success in *being respectful* is reflected in both the qualitative and quantitative findings. Not only did participants describe the curation of, and participation in, a respectful environment as key to their experience, but they also indicated agreement in more than 97% of responses (45/46) to both the survey statement of *feeling respected* and being *seen as an expert* during co-design workshops. Furthermore, one participant described new-found confidence in their stakeholder expertise after participating in P-POD. Increased stakeholder confidence has also been reported as a key benefit for participants during in-person co-design processes [8,13,57].

Many P-POD participants spoke very strongly regarding their feelings of leadership in decision-making and ownership over the final design. This aligns with the aim of P-POD to sit within the ’collaborate’ domain of the IAP2 Spectrum of Public Participation, where research involves *“partnering with stakeholders to identify preferred solutions, and incorporating their advice and recommendations into research decisions to the maximum extent possible”* [20]. Participants also appreciated attempts to power-share such as taking on tasks for facilitating the workshops, however, many described being satisfied with the amount of input they were giving, and not having the capacity or desire to take on further roles. As one participant noted, perhaps further directive support could be given for these roles, to ensure that participants truly can step into these confidently should they wish. However, as survey findings indicated that participants felt they “had enough support to contribute” (45/46 responses) perhaps participants were comfortable with the amount of power and responsibility they had. Although participants certainly felt agency in certain parts of the process, and they did feel ownership and desire to see it progress into the implementation stage, many participants felt that ultimately, responsibility for the process fell to ResearcherA due to their role as project lead role. Participants recognised that more responsibility equated to more burden. Some participants expressed that additional responsibility outside of scheduled workshops was beyond their capacity to invest in the project at the time. These findings indicate that to facilitate P-POD some team members (such as the project lead) must assume increased responsibility and complete extra tasks in-between group workshops. This prompted the authorship team to discuss and debate: should research teams strive for equality for all members throughout an online research co-design process? Based on the evaluation findings, the authors posit that power-sharing in P-POD should instead focus on equity and inclusion, rather than equality. As Farr [58] comments, power in co-design should be considered carefully, with the goal being that all stakeholders should benefit from its use, rather than subscribing to an ’ideal’ of equality. *“Collective power”* [58,p.628] derived from working *with* stakeholders should result in collaboration, respect, and connecting with participants’ values [58]. Participants reported these benefits in describing their experience in P-POD, and therefore moving into the ’empower’ domain of the IAP2 Spectrum where stakeholders completely drive the research may not always be the ’ideal’, particularly when working with *“time-poor”* (ClinicianB) parents and clinicians.

Pallesen et al., [10] also reflected on the co-design environment and power-sharing as key themes developed from their qualitative evaluation of participants’ experiences of a face-to-face research co-design process. Pallesen et al. reported that participants experienced a sharing of power via collaborative leadership in their co-design process which was actively facilitated by participants influencing the agenda and workshop direction. However, participants in Pallesen et al.’s study reported feelings of anxiety and uncertainty in the first few workshops [10]. In contrast, participants in P-POD reported the first couple of sessions as being particularly key to forming the respectful and participatory environment of P-POD. Potentially, the positive experience of the first sessions in P-POD may be due to the familiarity of the project and team members from their involvement in the prior study. This aligns with Pallesen et al.’s suggestion that the easing of their participants’ anxiety and uncertainty over time was due to building relationships and gaining familiarity with the processes [10]. Engaging participants in a prior stakeholder engagement project and careful design of initial co-design sessions may accelerate relationship building and confidence within co-design teams. Furthermore, these strategies are likely essential to forming the respectful culture that enabled participants to *“step forward”* (CoachA) into shared power and agency.

The guiding principle of *being inclusive* in P-POD is defined as bringing together and supporting the involvement of diverse stakeholder groups in multiple ways and making clear the value of all participants and contributions. In theme 2, participants highlighted the value of diverse perspectives amongst team members which they suggested enriched both the co-design process and the final intervention design. Appreciation of differing stakeholder perspectives and expertise is also highlighted in the literature on in-person co-design processes [2,57,59]. Unlike the findings from P-POD’s evaluation, Gustavsson and Andersson [59] reported an initial reluctance by clinician stakeholders to work with end-users for fear of criticism. Similarly, Hyett et al. [2] describe participants’ challenging each other’s professional roles. This was not reflected in the P-POD team, where clinicians valued the input of parents in challenging some of their assumptions, but they were not challenged in their stakeholder role or expertise. This may be due to the respectful and supportive culture that was actively curated in P-POD, as well as parents’ positive experiences in the healthcare system which formed part of their purpose for participating in the co-design process. Furthermore, undertaking the co-design via an online platform rather than a hospital or research institution setting may facilitate a more neutral environment that may provide less inherent reminders of traditional power hierarchies. In theme 3, P-POD team members also spoke strongly about appreciating the multiple methods available to support contributions during and between workshops, and for one participant, this made the difference between participating and not. Within the differing methods of contributing, and the diverse perspectives involved, all participants commented on feeling that their input was important and valued in both the qualitative and quantitative findings. Literature on in-person co-design reports that feeling heard and valued, and experiencing new and stronger social connections are key benefits for participants [8,13,57], which was reflected in P-POD’s evaluation. Palmer et al. [57] describe that feeling heard and valued within a co-design team contributes to a shared purpose and a sense of *“collective identity”* [57,p. 253] which motivates the group to connect past experiences towards creating future solutions. P-POD participants also reported a sense of shared purpose, and appreciated the small group discussions in breakout rooms as facilitating the building of relationships (or collective identity) between the team members. However, some participants wondered if a hybrid model for those who could attend face-to-face might further cement developing relationships. Fails et al. [24] and Kennedy et al. [25] both point out that hybrid models have the potential to act unfavourably on team dynamics, with those joining virtually feeling like *“outsiders”* [24.p.3] compared with the in-person members.

The principle of *being participatory* is defined in P-POD as ensuring participants have the skills required to participate as much as they want to, in the way they want to. In theme 4 and survey findings, participants described obtaining knowledge and skills in co-design and the use of online collaborative platforms. As described above, team members felt they had agency in their participation, and this is supported by 98% (45/46) of responses agreeing with the survey statement “I could participate if/ when I wanted to”. Participants discussed the lack of judgement from other team members during sessions and appreciated the room to debate differing opinions. Pallesen et al. described their face-to-face co-design process as having an *“absence of conflict or disagreement”* [10,p.363], whereas P-POD seemed to provide an environment where participants felt safe to share and debate diverse perspectives, and working toward consensus was a valuable part of the process. Strategies to support and encourage space for healthy debate within P-POD were setting collaborative ground rules, and the use of tools such as the Consensus Decision-Making framework and Nominal Group Technique.

The final guiding principle of P-POD is *being outcomes-focused.* P-POD defines this principle as continually working towards the clearly defined objective/s of the process as well as participants’ individual goals. In theme 1, participants reflected on the benefit of understanding the project’s vision before starting the co-design process, and how this was helpful to keep discussions on track during the workshops. All participants felt that the process had achieved the project goal, but many felt that implementing the rehabilitation intervention was a critical part of the process (theme 4). A benefit reported in the co-design literature is participants gain knowledge and expertise as part of their involvement [2,8,13]. P-POD team members reported gaining knowledge that they would utilise beyond the project, such as an increased understanding of co-design, consensus decision-making, and circus activities.

By mapping the evaluation results to guiding principles of the process, and contextualising within contemporary literature on co-design approaches, it is clear that the experiences and benefits reported by P-POD team members align with those described by in-person research co-design approaches. This provides emerging evidence that P-POD is a process that results in an authentic translation of research co-design into an online environment. Key P-POD strategies valued by participants to uphold guiding principles are outlined in Figure 6. Additional strategies undertaken by the research team can be viewed in Table 1. These strategies can be implemented in future research co-design in an online environment.

**Figure 6:**
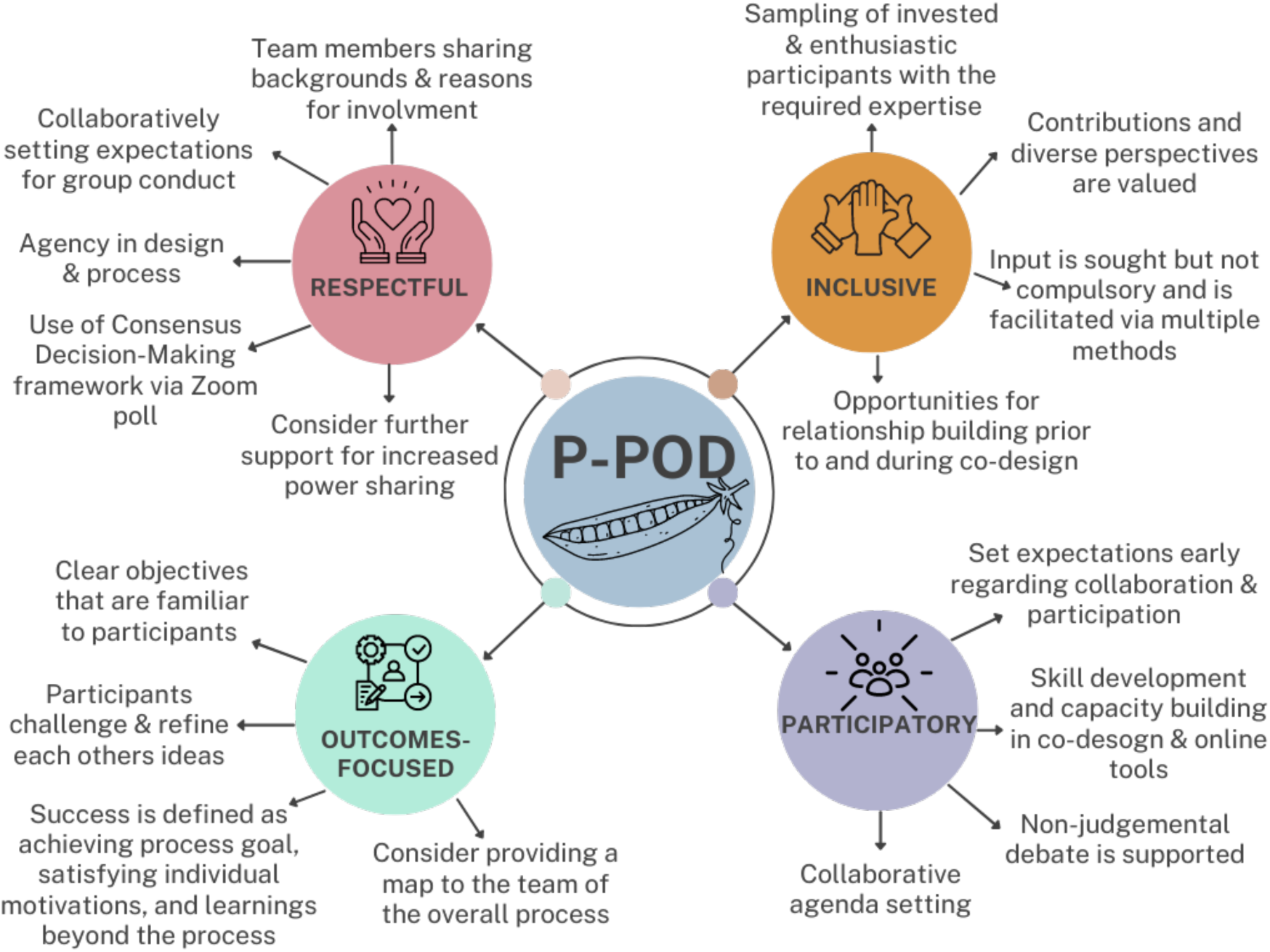
P-POD strategies valued by participants. Figure 6 Alt Text: A mind map showing a centre circle titled P-POD, with four branches titled respectful, inclusive, participatory and outcomes-focussed respectively which list strategies valued by participants.

### Strengths and limitations

This study adds to the limited evidence base regarding the evaluation of online co-design approaches and provides clear recommendations for future co-design processes aiming to authentically transition to an online environment.

There is a possibility that P-POD team members participating in evaluation interviews felt pressured to speak positively about their experiences due to social desirability bias. However, having an interviewer unknown to the participants who had not been involved in P-POD was a strength of the evaluation, and the primary strategy used to address this issue. Furthermore, one participant commented on appreciating the opportunity to reflect on their experiences *“knowing that I could say bad things too. But there was no bad things”* (ParentB). Confirmation bias may have affected the analysis of the qualitative evaluation data, and the primary strategies used to address this was using a collaborative approach with three analysts, as well as member-checking the results with participants. No changes to thematic findings were requested by participants during member checking.

The duration of participant involvement in P-POD was approximately 12 hours which is shorter than the 18-28 hours recommended by Leask et al. [17]. However, the work done in a prior study with the same stakeholders [44] may have resulted in an accelerated process as participants where already aware of the project topic and key considerations. Furthermore, the co-design team did not include children with lived experience of being born preterm due to the age range of the intended intervention, and the assumption that parents would be best placed to inform interventions for this age group (3-5 years). However, future research should consider involving older children or adults with this lived experience to further enrich the intervention design.

Although this research aimed to ’collaborate’ and not ’empower’ on the IAP2 Spectrum [20], in future research the authors would recommend including stakeholder input even earlier (e.g. setting the initial research question). This strategy may further address power imbalances, without placing an undue burden on stakeholders. As this current study formed part of F.C.’s doctoral research, the research question and focus needed to be decided before the co-design team was engaged. This issue was also noted in other co-design research but was not considered detrimental to the process or outcome [2].

Although the evaluation described in this article is primarily process-based, formal outcome evaluation of the co-designed intervention is planned via a pilot study. The literature on research co-design also describes additional components considered in outcome evaluation which were addressed in the current study. These include participant self-reported measures of personal development and learning [21], evidence that stakeholder expertise has influenced the outcomes of the project [21,59], and results demonstrating that the outputs of the codesign are effective in addressing research questions [2]. Furthermore, co-design team members continue to be involved with both the co-production of the intervention and the pilot study.

## Conclusion

P-POD is a novel online research co-design approach that aligns with principles from the literature on co-design and stakeholder involvement. P-POD is respectful, inclusive, participatory, and outcomes-focused and appears to impact the lives of stakeholders involved in the process in a positive and meaningful way. Overall, participants reported engagement and satisfaction with the co-design workshops. They also perceived that P-POD adhered to its four guiding principles. The experiences of the co-design team members articulated the benefits of P-POD which align with those reported in the literature regarding in-person co-design approaches. This may indicate that P-POD is a process that results in an authentic transition of co-design into an online environment. Furthermore, P-POD can be used to collaboratively design paediatric rehabilitation interventions with key stakeholders while providing increased accessibility for stakeholders to be involved in research that affects them.

## Data Availability

The data that support the findings of this study are available from the corresponding author, FC, upon reasonable request.

## Acknowledgments

We would like to thank the families of the Victorian Infant Collaborative Study (VICS 2016-2017) for their participation in this research, and the VICS research team, in particular Joy Olsen and Marion McDonald. Thanks to the Victorian Infant Brain Studies statistician Rheanna Mainzer for their support with the quantitative data analysis. This study will contribute to F.C.’s PhD candidature through The University of Melbourne.

## Appendix 1: Evaluation Survey & Interview Question Guides

### Evaluation of co-design process

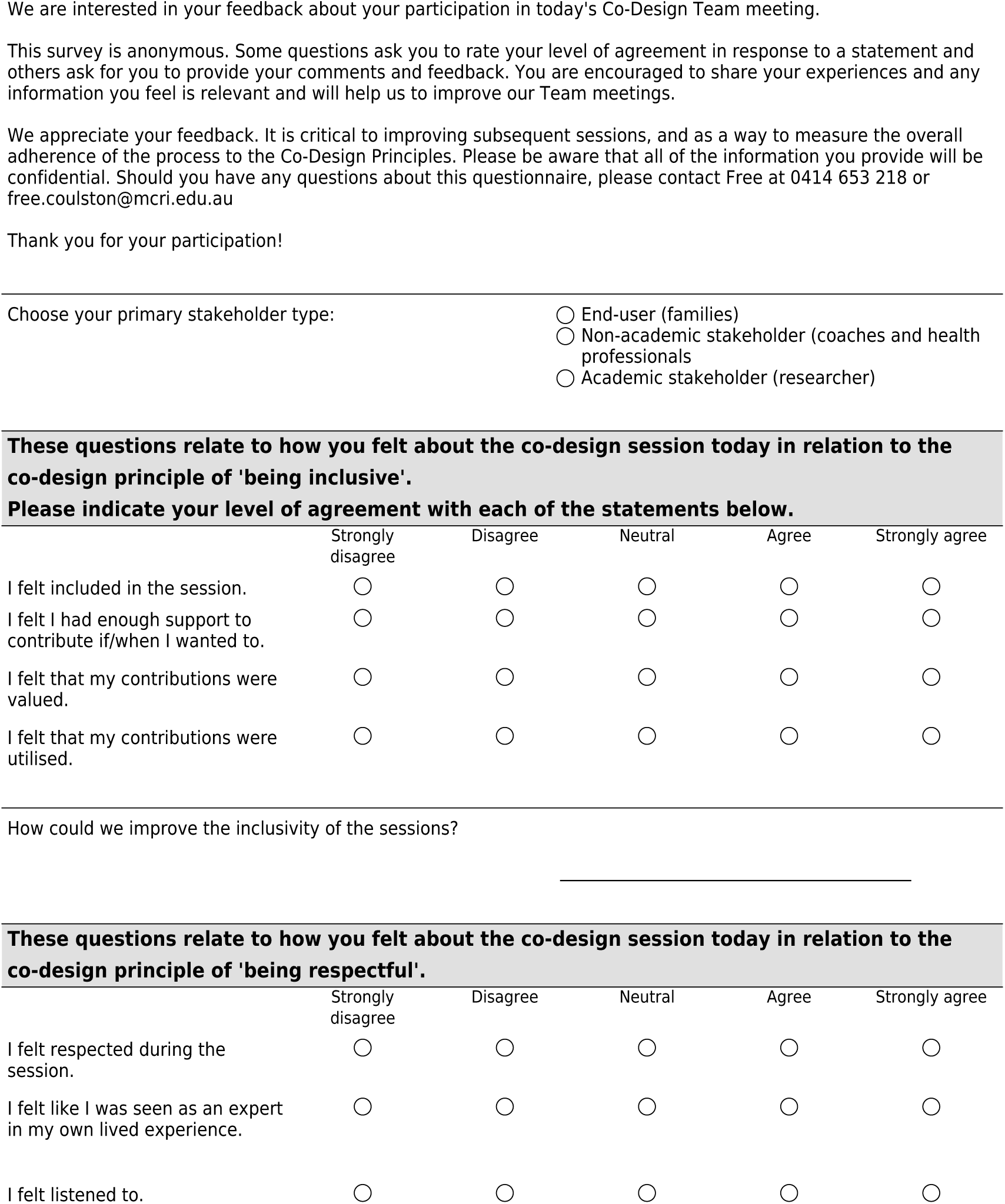

#### Interview Guidelines for Co-Design Participants

Adapted from questions utilised in the literature [5,8,10,17,37,45,60].

Please note: this is a GUIDE only – this is a semi-structured interview design so please modify as needed to respond to participants, or as dictated by your expertise.

##### WELCOME AND INTRODUCTIONS

**Aim:** To make participants feel at ease and ensure they are clear about what to expect and the aims of the interview

- Thank them for giving up their time to speak with us (make them feel welcomed and valued)
- Introductions
  - *My name is Kate Cameron, I am assisting with the evaluation of the co-design process you have just been a part of, and I will be asking you some questions about your experiences*
- Provide a very brief description of the research due to pre-existing knowledge of the project & explain how we will use their insights
  - *It’s important to evaluate the process of co-design from the participants’ perspective to understand the benefits and challenges of this method, where improvements can be made, and how it affects those who use it*.
- Explain how the interview will work (format and length):
  - *The interview will take approximately 30-45 minutes*.
  - *I can repeat or clarify questions, if I have been unclear, however you can choose to not answer certain questions, just let me know and I will move on*.
  - *There are no right or wrong or silly answers, I would like you to answer as richly and fully as possible, and I will ask some follow up questions as we go to assist this*.
  - *You can take a break or stop at any time, please just let me know*.
- Re-affirm consent verbally, allow for any questions before asking Interview Questions.
  - *Do you mind if I record this interview so that I can concentrate on what you are saying? PRESS RECORD*
  - *Did you have any further questions before we get started?*
  - *Do you voluntarily consent to continue with the interview?*

##### INTERVIEW QUESTIONS

**Aim:** To gather insights that will help achieve the study aims.

<u>Prompting and probing</u> will be asked based on the participants’ response. Before moving to a new topic, the interviewer will <u>summarise the main points</u> of the participants’ answer to check they are representing their intention correctly.

*1. Now, Free has told me that you have/ haven’t participated in a co-design project before*.
  *1.1. Have (IF only):*
    *1.1.1. Can you describe your understanding of co-design for me?*
    *1.1.2. How did The Co-Design Team’s process compare to your previous experience?*
    *1.1.3. How do you think an online context impacted the co-design process?*
  *1.2. Haven’t (all others):*
    *1.2.1. Can you describe your understanding of co-design for me?*
    *1.2.2. How do you think an online context impacted the co-design process?*
*2. Can you describe your experience of the co-design process for this project, from the initial survey, interview, focus group (parents only) through to the Co-design Team?*
  - *Probing questions (if needed)*
    - *How did it make you feel? What is an example of the best part? What about the worst part?*

*Thinking now specifically about The Co-Design Team Meetings:*

*3. How do you feel about what the Co-Design Team achieved in regards to it’s goal:* to co-design an intervention using circus activities that improves physical activity participation for pre-schoolers born preterm, using the information collected from parents, health professionals & circus coaches, and the combined expertise in the Co-design Team.
  - *Probing questions (if needed)*
    - *Would you consider the process a success? Can you elaborate a little on that?*
*4. In the first session, you were asked what you were hoping to get out of this process and you described….*.
  - *Participant 1: wanting to learn more about co-design and network with likeminded people*
  - *Participant 2: wanting to ‘give back’ as a result of the care you and your boys received*
  - *Participant 3: interest in research and keen to contribute to the development of a program which is evidence-based and could impact policy*
  - *Participant 4: wanting to create a program which is appropriate and inclusive for kids born preterm and offers parents a chance to connect*
  - *Participant 5: seeing a lot of potential for this project to make a difference in the community*
  - *Participant 6: hoping to create a program that would benefit future clients*
  - *Participant 7: wanting to assist in creating a program that would improve outcomes for children born preterm*
  - *Participant 8: wanting to work with and learn from the team, hoping to implement the circus program we codesign*.
*4.1. Can you describe how this may or may not have occurred?*
*4.2. What other expectations did you have of the co-design process?*
*5. What do you think worked well about the co-design process?*
  - *Probing questions (if needed):*
    - *What was the most satisfying or enjoyable experience? What were some of the advantages to participating in this co-design project? What did aspects did you enjoy/find interesting or useful?*
*6. What do you think did not work so well about the co-design process?*
  - *Probing questions (if needed):*
    - *What was the most frustrating or unenjoyable experience? What were the challenges/barriers and what (if any) steps were taken to overcome these challenges?*
    - *Do you think everyone was engaged in the workshops? Do you think this changed over time? If not, what do you think might have prevented people from engaging with the workshops?*
    - *What should we do differently next time we use co-design?*
*7. Has anything changed in the way you live, work or think about things as a result of participating in this project?*
  - *Probing questions (if needed):*
    - *Did being involved in the project have any impact on you, whether positive or negative?*
    - *New knowledge, new skills, new networks, new understandings, new opportunities, new possibilities?*
*8. How likely is it that you would participate in another project that uses co-design after this experience?*
  - *Probing questions (if needed):*
    - *Can you tell me a little more about that?*
*9. Is there anything else you would like to add about your experience that I haven’t asked about?*

##### WRAP UP

**Aim:** To make consumers feel valued and give them clarity on what will happen next

- *Thank you so much for speaking with me today, Free will be in touch with further information about the next steps for this project*.
- *Would you like to receive a written copy of this interview to check that it reflects your experiences the way you intended?*
- *Would you like to see the de-identified analysis of all of the interviews so that you can make sure that it resonates with your experiences?*
- Thank them and sign off

## Appendix 2: TIDieR Checklist for CirqAll: Preschool Circus for Premmies (adapted from Hoffman et al. **[52]**)

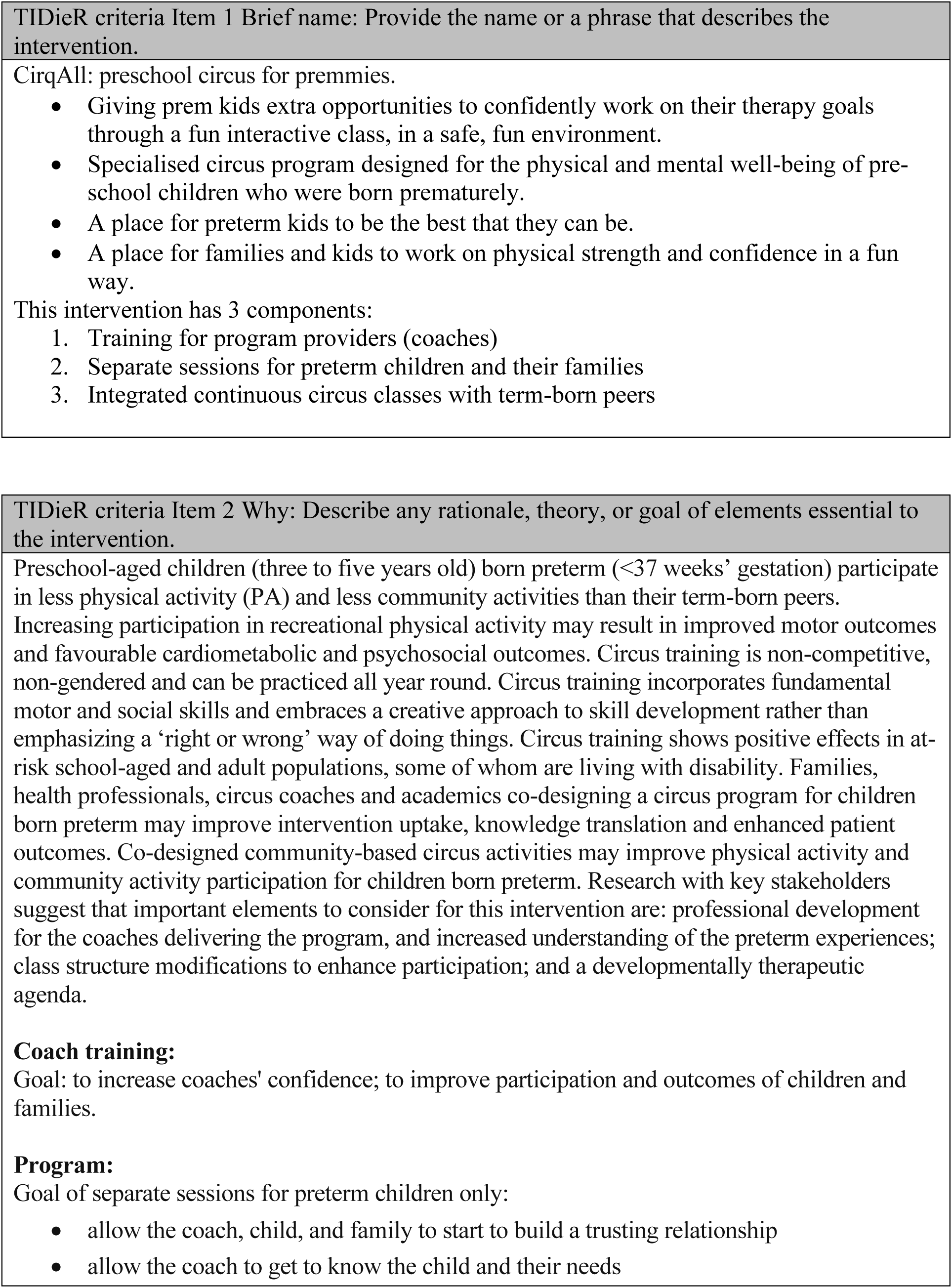
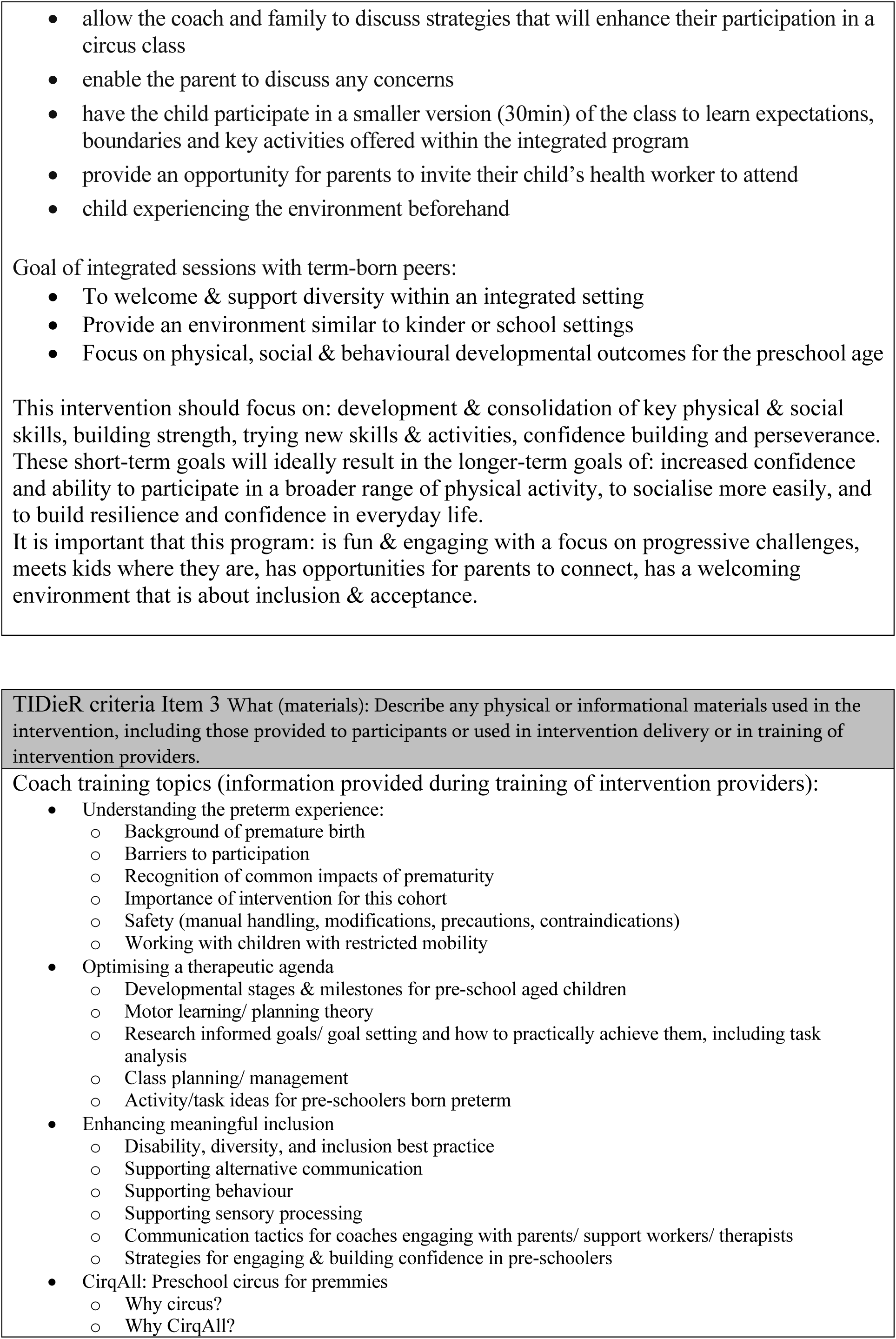
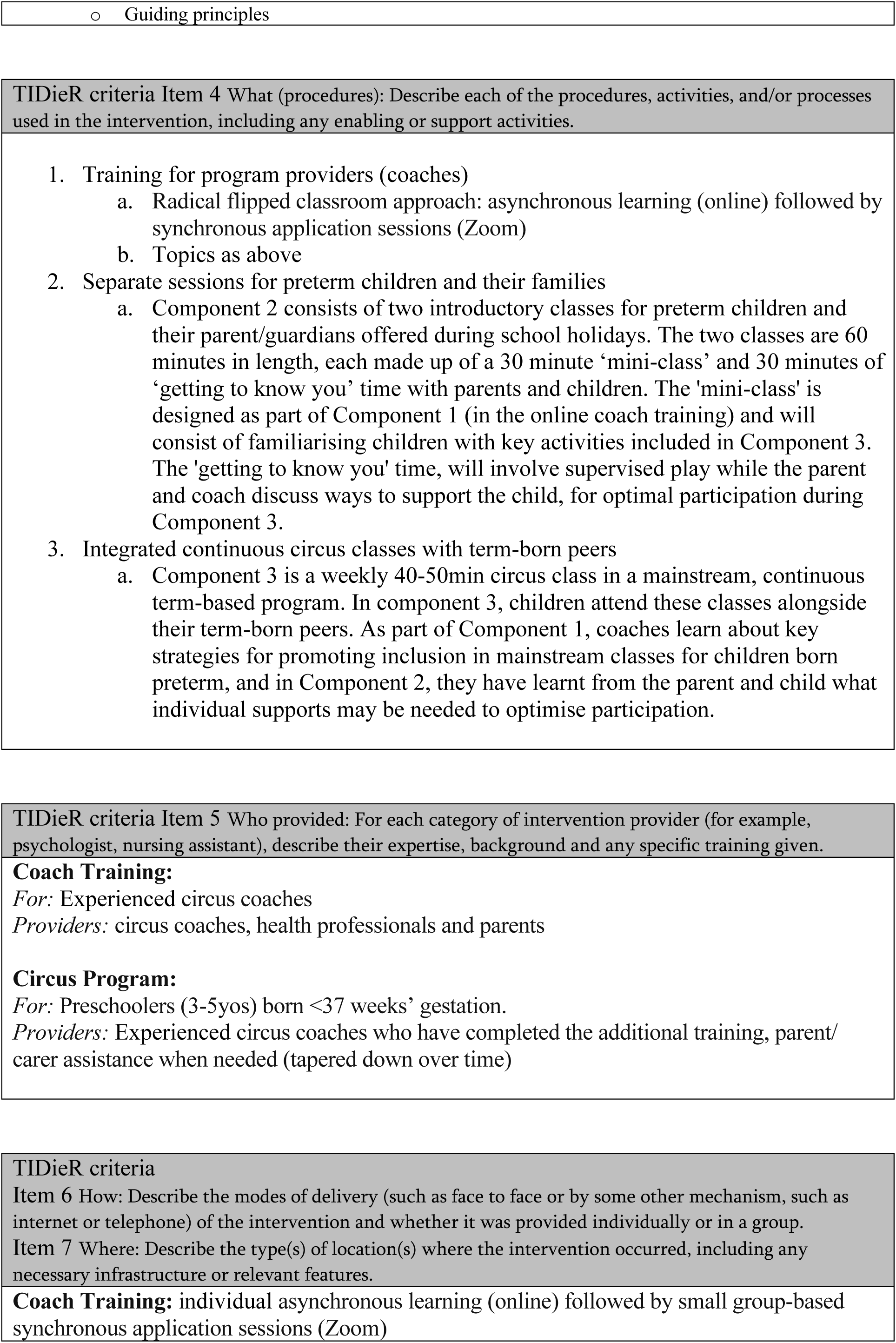
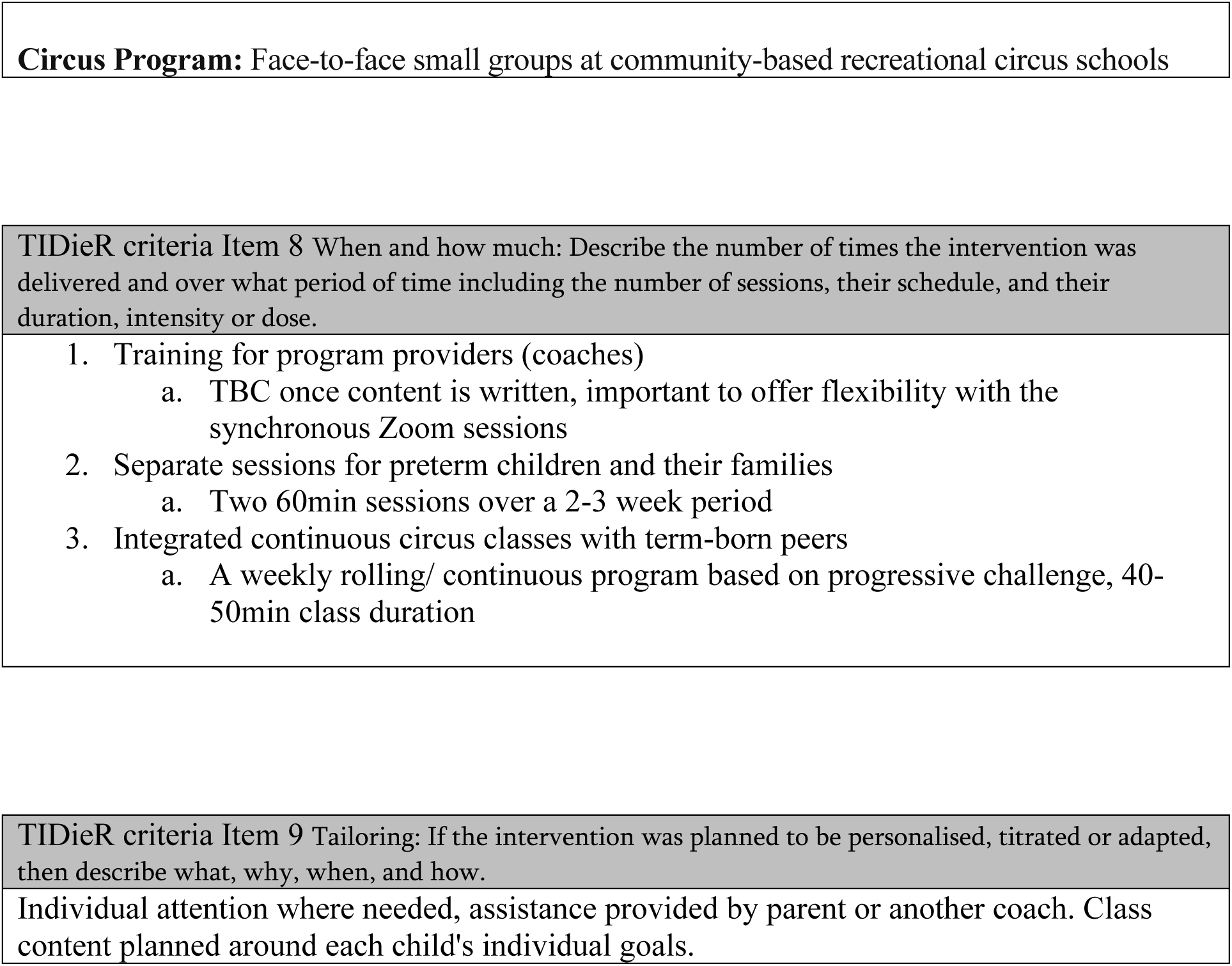

## Appendix 3: Project lead’s detailed reflexive statement

The project lead and primary analyst (F.C.) maintained their reflexive practice throughout the co-design and analysis phases of this research by journaling, participating in an evaluation interview, fortnightly meetings with PhD supervisors (A.S., R.T. and K.S.), and meetings with the co-analysts (C.M. and K.C). This reflexive statement draws on all these experiences to present an overview of the values and assumptions that F.C. bought to this research and the interpretation of the findings and is thus written in first person.

This research sits in a constructivist ontology and epistemology. As both a feminist and a physiotherapist, allowing space for diverse stakeholder voices, and particularly those traditionally not given power in health research and decision-making, was extremely important to me throughout this research. I identified with three of the four different stakeholder groups in the co-design: I am a circus coach, a clinician, and a researcher, and although I am a mother, I have never parented a child born preterm. Creating a space where these parents could feel comfortable and valued and important was so fundamentally essential to me. However, having strong values of truth-seeking and desire for clarity (both personally and in my clinical work), meant I needed to consistently reflect on whether I was continuing to be open to multiple realities and resisting a positivist, objective lens. Certainly, over my qualitative journey as a researcher I have become much more comfortable sitting in the discomfort of remaining open to change throughout the analytic process in order to produce a strong, complex and thoughtful interpretation. Becoming comfortable with discomfort is a key learning I took into the co-design process and continued to learn from in this space. Slowing down, and valuing the process and relationships as much as the outcome was another key lesson I took away from the co-design, and continue to implement in my personal and professional life.

My assumption going into the co-design process was that we wouldn’t be able to please everyone with the end design, and that people would be unhappy if they needed to compromise. I think my biggest learning in this space is that with transparent and truly collaborative decision-making processes, and people’s respect for each other’s expertise (lived or professional), that the end design is not a product of compromises, but rather a product of each person’s knowledge, creativity, and adaptability. This has changed my career path as a researcher, as I now actively seek out collaborative work as I value it so highly.

The experience that I was unprepared for in P-POD, which also shapes my future practice, was the extent of the emotional labour required to project lead and facilitate this process. I felt a huge responsibility to the team members to make sure that each participant felt included and involved in a way that made them feel capable, satisfied, and cared for. I also felt a huge responsibility to the children and families that would be the end-users of this program. This sense of responsibility meant that I bought absolutely everything I had to this project, both in the planning and implementation and the work between sessions, leaving very little emotional energy for other aspects of my personal and professional life over that time. I think it’s important to plan for this emotional labour when considering engaging in co-design and include additional plans for your self-care as project lead and/or facilitator to ensure the sustainability of your emotional well-being throughout the process.

My assumptions going into the evaluation data analysis were that the co-design team members were going to be very critical of the process, and to have found it overwhelming and exhausting. As I had quite literally spent six months of my life trying to design a process that would be authentic and inclusive, I felt very sensitive to criticism and feelings of failure, so it took me a number of months to be brave enough to engage in the evaluation analysis due to my fear of negative feedback. Once I engaged with the data however, I quickly realised that this was not the primary experience of the team, and in fact, in listening to my own interview data, I was the only participant that strongly articulated these feelings. For this reason, choosing a collaborative approach to the analysis was essential to keep a sense of openness to the data, and to be able to work through any constructive criticism in a sensitive and future-focussed way.

